# Evaluating targeted COVID-19 vaccination strategies with agent-based modeling

**DOI:** 10.1101/2023.03.09.23285319

**Authors:** Thomas J. Hladish, Alexander N. Pillai, Carl A. B. Pearson, Kok Ben Toh, Andrea Tamayo, Arlin Stoltzfus, Ira M. Longini

## Abstract

We evaluate approaches to vaccine distribution using an agent-based model of human activity and COVID-19 transmission calibrated to detailed trends in cases, hospitalizations, deaths, seroprevalence, and vaccine breakthrough infections in Florida, USA. We compare the incremental effectiveness for four different distribution strategies at four different levels of vaccine availability, reflecting different income settings’ historical COVID-19 vaccine distribution. Our analysis indicates that the best strategy to reduce severe outcomes is to actively target high disease-risk individuals. This was true in every scenario, although the advantage was greatest for the middle-income-country availability assumptions, and relatively modest compared to a simple mass vaccination approach for rapid, high levels of vaccine availability. Ring vaccination, while generally the most effective strategy for reducing infections, ultimately proved least effective at preventing deaths. We also consider using age group as a practical, surrogate measure for actual disease-risk targeting; this approach still outperforms both simple mass distribution and ring vaccination.

We also find that the magnitude of strategy effectiveness depends on when assessment occurs (*e.g*., after delta vs. after omicron variants). However, these differences in absolute benefit for the strategies do not change the ranking of their performance at preventing severe outcomes across vaccine availability assumptions.

## Introduction

During outbreak or pandemic situations, public health agencies respond with various interventions to contain and mitigate spread of the pathogen. When available, vaccination can be a useful strategy to reduce both transmission and infection severity. However, different deployment strategies for vaccination vary in effectiveness, and in ways that may depend on vaccine performance and the natural history of the infection (*1–3*). During the COVID-19 pandemic in the United States (USA), vaccination generally proceeded via passive, mass distribution, from older to younger age groups, with priority access for *e.g*., health care workers and individuals at high risk for severe outcomes (*4*). However, vaccines can also be used in more proactive strategies, like ring vaccination for Ebola, where responders vaccinate contacts and contacts-of-contacts of identified cases (*5*).

During outbreaks, vaccination-as-containment strategies are often reserved for pathogens with a relatively long generation time, so that vaccination can be administered and achieve efficacy prior to exposure (*6, 7*). Essentially, the vaccination effort needs to outpace the spread of the pathogen. For pathogens that spread very quickly or frequently cause infections without a known exposure, vaccination strategies aim to vaccinate the most people, often preferring those with higher risk for severe outcomes (*8, 9*). In general, “passive” interventions—where officials issue guidance, and make resources publicly available—will tend to be less resource-intensive than “active” interventions that require seeking out intervention targets. Choice of strategy may also depend on vaccine performance: containment requires efficacy against transmission, whereas targeting high-risk individuals requires a vaccine that provides good protection against severe outcomes if infection occurs.

SARS-like (*e.g*., SARS-CoV-1, MERS) outbreaks have been controllable in the past with a range of active measures (*10, 11*). In some settings, SARS-CoV-2 responses have included substantial, effective contact tracing programs while infection prevalence was relatively low (*12, 13*). Plausibly, active vaccination approaches would be beneficial as well. In this study, we evaluated various vaccination strategies using a detailed agent-based simulation model of human activity and SARS-CoV-2 transmission. With reference to a counterfactual simulation where vaccines are distributed passively (*i.e*., via a proportional-allocation mass campaign), we evaluated the effectiveness of active strategies, including quarantining, ring vaccination, age-prioritized vaccination, and true risk-prioritized vaccination. Many locales have prioritized vaccination by age, as a proxy for knowing actual, individual-specific risk of hospitalization (or death) given infection. As our simulated risk-prioritization strategy has this information, we consider it to be an upper bound on the performance of this kind of strategy.

We further evaluated whether strategy ranking is sensitive to vaccine availability by considering four levels of availability, including three chosen to represent low-, middle- or high-income countries world-wide (hereafter LIC, MIC, and HIC respectively). We also evaluated strategies based on data specifically for the USA, which had particularly fast early uptake of the vaccine, followed by slower-than-HIC uptake during the second half of 2021 (*14*).

## Methods

We extended an agent-based model framework to support evaluating the COVID-19 epidemic and response efforts (Fig 1), previously used in (*15*) and derived from (*1*). For a detailed description of the model, see S1 Additional Methods. The model represents SARS-CoV-2 natural history with empirically-based mechanisms in a stochastic, discrete-time transmission simulation of people moving between places (Fig 2). For this analysis, we consider a population of 375K people with demographics representative of Florida (and spatial structure corresponding to Marion County, Florida) as a practical sub-population with both urban and rural areas. We sample the 162K households in the model from state-wide microcensus data, with features including household size, ages in years, and employment or student status, and validate the model against state-wide data (Fig 3). The presence of comorbidities relevant to COVID-19 is known in aggregate but not at the household level, and thus is sampled independently. Locations in the model (aside from residences) are based on their actual addresses in Marion County, including 46 long-term care facilities, 33.8K workplaces, 118 schools, and 6 hospitals.

**Figure 1:**
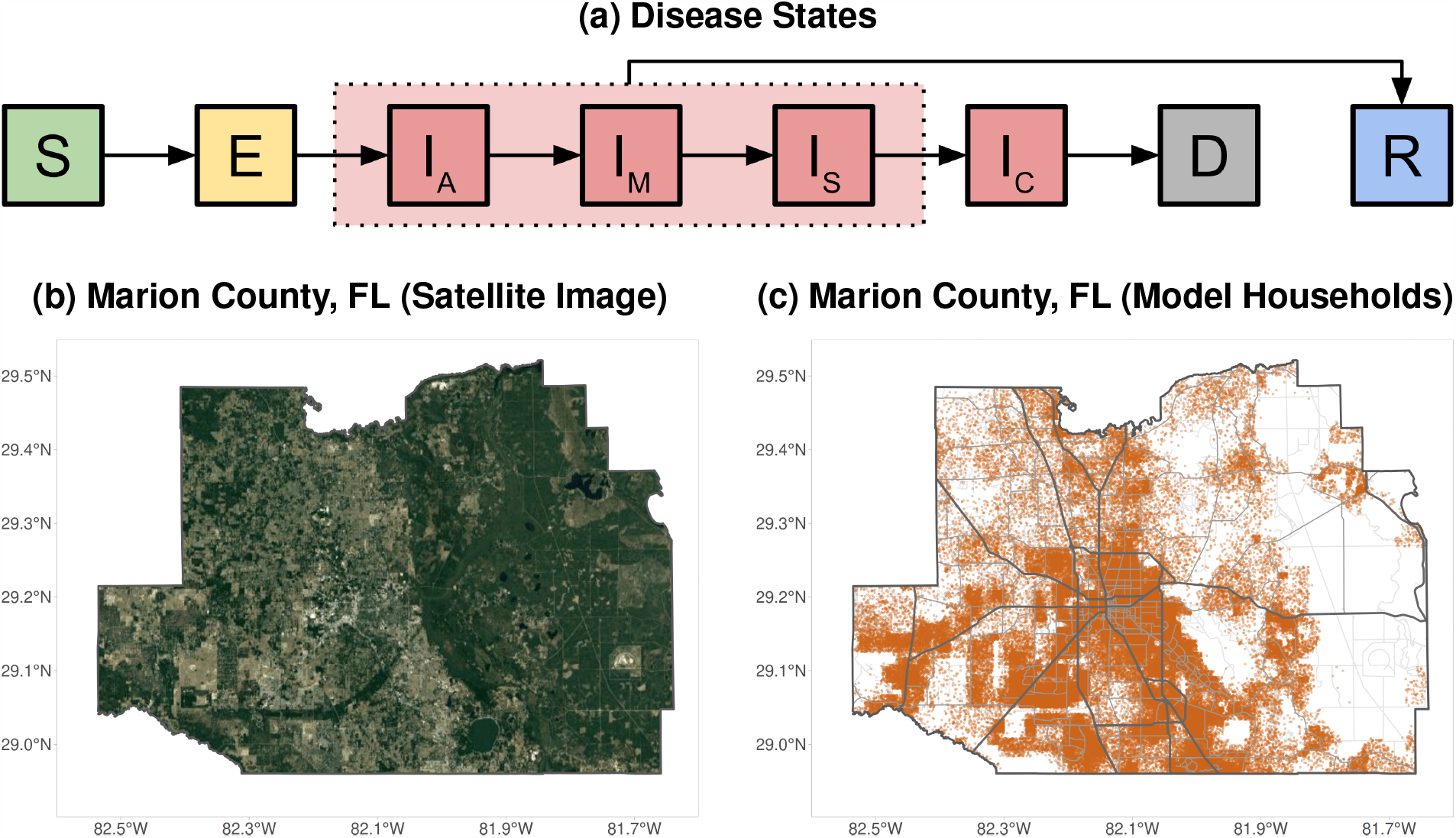
Model disease states and spatial structure. (a) Progression of the disease states in the model: susceptible (*S*) individuals may become exposed (*E*) to the virus, then progress to being infectious (initially asymptomatic [*I*_*A*_], possibly progressing to mild [*I*_*M*_], severe [*I*_*S*_] or critical [*I*_*C*_]), and finally recovering (*R*) or dying (*D*). Recovered individuals have strain-specific immunity that changes over time. (b) Satellite image of Marion County, FL, the region used for the model’s spatial structure. (c) Locations of the 115K model households (orange dots). Roads are shown for reference but are not modeled.

**Figure 2:**
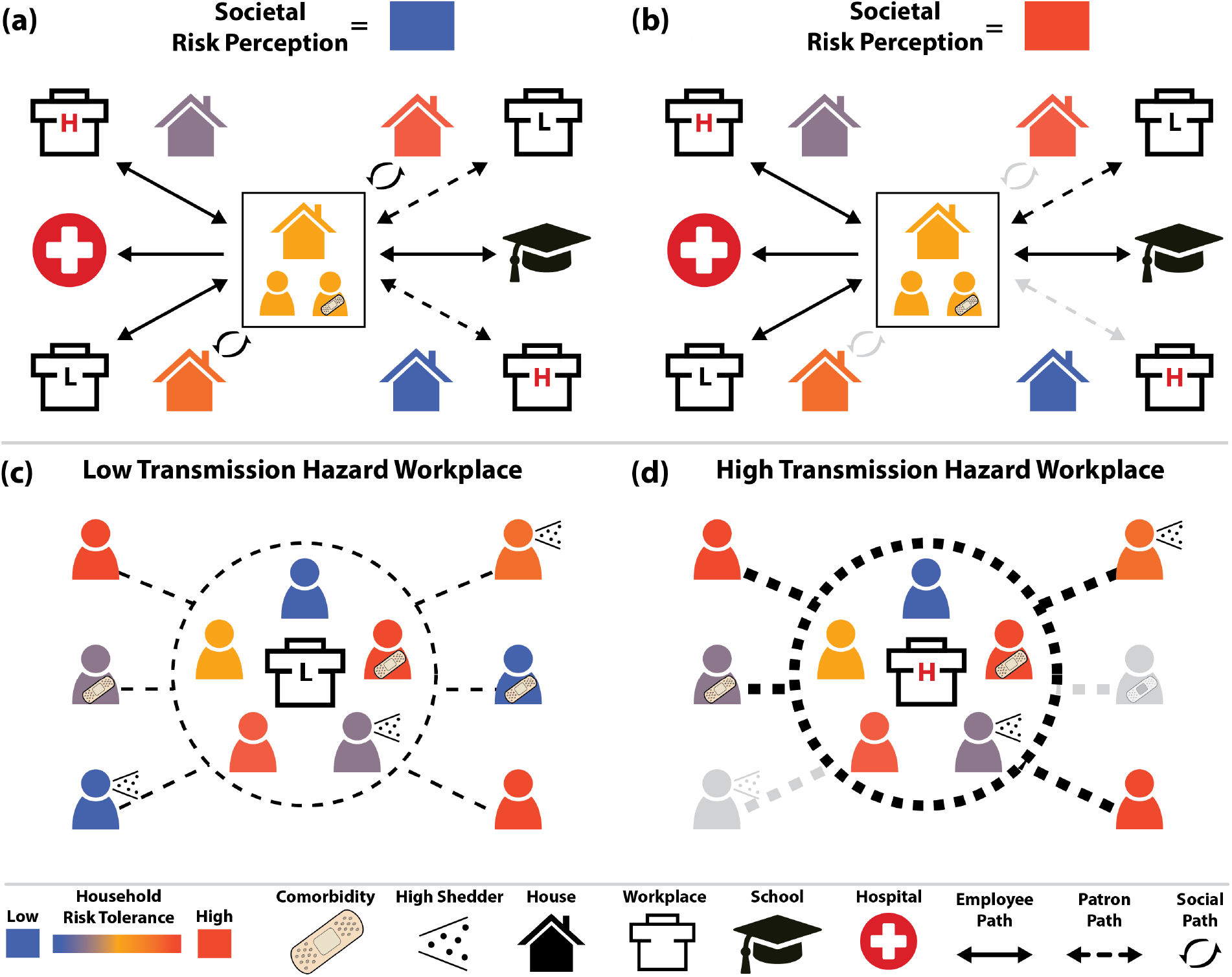
Individual interactions and behaviors in the model. Interactions occur when people in the model are in the same location at the same time, and may occur in households, workplaces, schools, hospitals, and long-term care facilities (not shown). Households have an inherent risk tolerance (indicated by color), and probabilistically have inter-household connections homophilously based on that tolerance. The population overall has a time-varying perception of risk of COVID-19 infection that may be different from the actual risk. (a) When the societal perception of risk is lower than a household’s risk tolerance, household members engage in all their normal activities, including socializing with specific other households and patronizing specific high-transmission-risk workplaces like restaurants and bars. (b) When the societal perception of risk exceeds a given household’s risk tolerance, the household will cease high-risk activities (indicated with greyed arrows), while maintaining more essential activities like going to work, school, and patronizing low risk workplaces (*e.g*., grocery stores). (c, d) Employees and patrons interact in some workplace types, with interactions between employees more likely to result in transmission. (d) When perceived risk is high, risk-intolerant (blue) employees of high-transmission-hazard workplaces still go to work, while risk-intolerant patrons cease their patronage.

**Figure 3:**
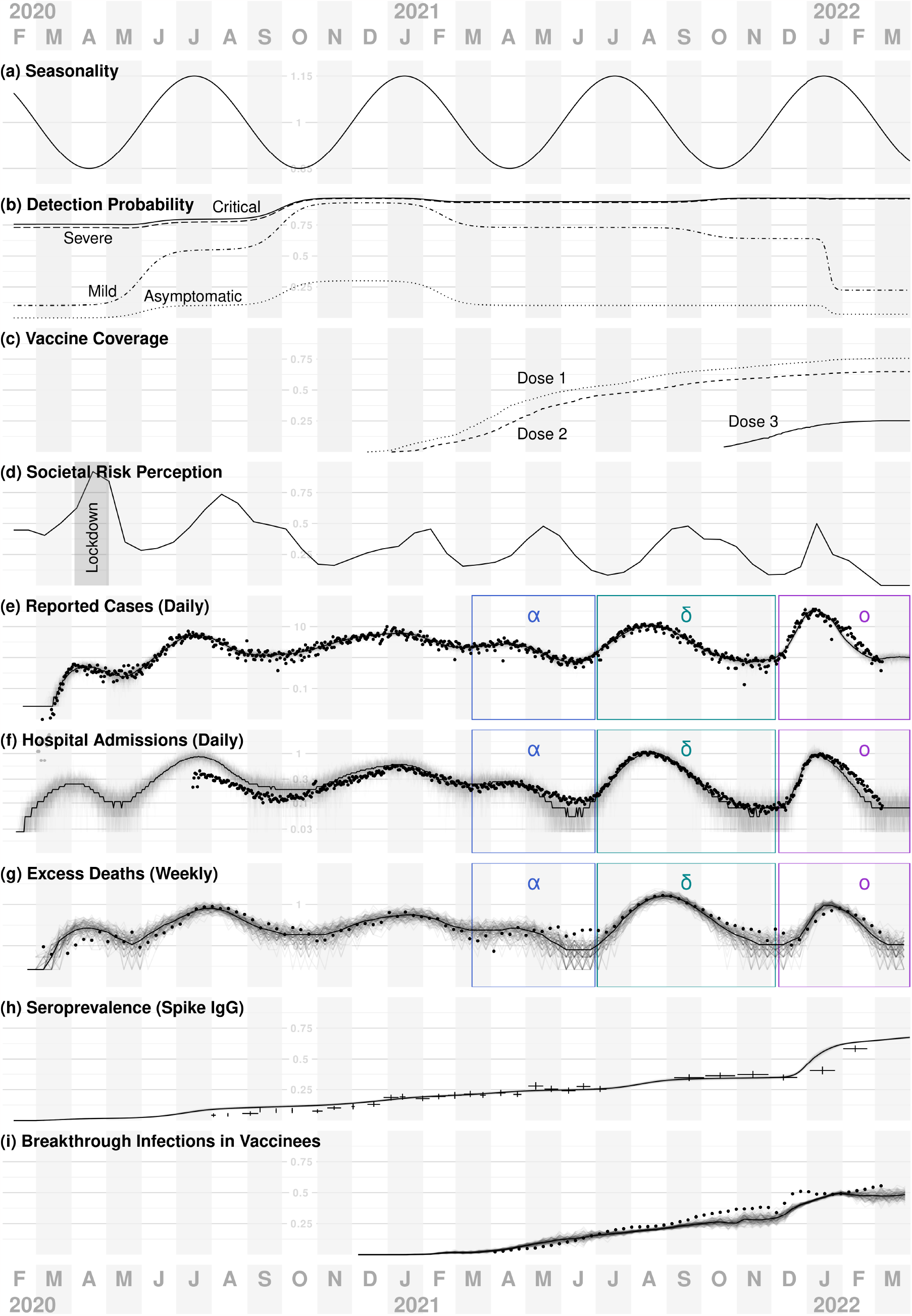
Time-varying model inputs and indicators of model performance. Panels (a-d) show model inputs, and (e-i) compare model outputs to observed data. In (e-i), points and cross-hairs indicate observed values, solid lines median trends, and faded lines sample trajectories. Horizontal gridline values are plotted above October 2020. (a) Seasonal forcing has a 6-month period, peaking in January and July each year. (b) Detection and reporting probabilities by disease outcome. (c) Simulated first, second and third vaccine doses distributed statewide in Florida, used to calibrate the model (but not for evaluating strategies). (d) Societal risk perception, which drives personal protective behaviors in the model, is fitted so that cumulative reported cases in the model match empirical case data for FL (black dots in panel e). For approximately the month of April 2020, non-essential businesses were closed in the state, and thus are closed during this period in the model (gray “lockdown” shaded region). Not shown: schools in the model close during the summers and during spring 2020, and are 50% and 80% open during the 2020-2021 and 2021-2022 school years, respectively. (e–i) Simulated data closely track empirical data for incidence of reported cases (e), daily hospital admissions (f), excess deaths (g), seroprevalence (h), and the fraction of infections that occurred in vaccinees (i). Results in (e—g) are scaled to show values per 10,000 individuals, and VOC waves are labeled as alpha (*α*), delta (*δ*) and omicron (*o*). For empirical seroprevalance data in (h), horizontal bars indicate the dates covered by each data point and vertical lines indicate the 95% CI).

People in the model engage in various interactions that may result in transmission and which are moderated by individual behavior thresholds (Fig 2). Some interactions always occur, like those within households, hospitals, and long-term care facilities. We model two population-wide non-pharmaceutical interventions (NPIs): “lockdown”, where only essential workplaces are open, and reduced activity in schools. These have specific periods and levels; see Section 5.3 in S1 Additional Methods. These NPIs apply in both the validation and scenario analysis simulations.

We also model personal protective behaviors (PPBs) which potentially moderate social interactions and patronage of businesses. PPBs are represented as individuals choosing not to visit social contacts and patronize high-transmission-hazard businesses if the societal risk perception exceeds an individual’s risk tolerance. Risk tolerance is static and is defined at the household scale, whereas societal risk perception varies by day but is the same for all households. See Section 5.4 in S1 Additional Methods for more details.

We represent the natural history of infection with exposed and infectious states, followed by a recovered state with and strain-specific immune memory (Fig 1a). Individuals vary in the efficacy of their immune responses to infection. Exposure and infection outcomes in the model are affected by age and immune history (see Section 1 in S1 Additional Methods for more details), and infection outcome is additionally affected by the presence of comorbidities.

The simulation advances in daily increments, with a series of transmission opportunities among individuals when they are co-located during their daily activity pattern (Fig 2). Age- and spatially-structured interaction patterns can emerge because activities vary by age and day of week, *e.g*., children attend specific schools, where they interact with other children from the model’s catchment area associated with that school, and individuals have specific businesses that they randomly patronize. For social interactions, households are more likely to be connected to other households with similar risk tolerance thresholds, thus infection risk-groups can emerge.

### Scenarios

We define model scenarios by three factors: (1) vaccine supply level (four levels), (2) vaccination distribution strategy (four strategies), and (3) quarantine policy (two alternatives). S2 Additional Results also covers a fourth dimension: whether eligibility to receive vaccine is conditional on not having a known prior infection, but the main text results only consider unconditional eligibility. The four supply levels represent LICs, MICs, HICs, and the USA (Fig 4). We express supply levels as available doses per 10K eligible people, and derive them from UNICEF estimates of vaccine deliveries (*14*) and WorldPop program estimates of population sizes (*16*). Doses administered on a given day are the lesser of doses available and number of eligible vaccinees under the vaccination strategy. Available doses that are not consumed are rolled over to the next day. As the age- and risk-based strategies never run out of eligible vaccinees, doses administered for those strategies match doses delivered as specified by UNICEF. The supply is distributed according to a vaccination strategy, either a passive mass vaccination “standard” program (*i.e*., completely random distribution) that we use as a baseline reference, or one of three active strategies, called “ring” (vaccinating some primary contacts of contact-traced cases, and some secondary contacts of contacts), “risk” (prioritized distribution by disease-risk deciles), or “age” (prioritized distribution by age-group deciles).

**Figure 4:**
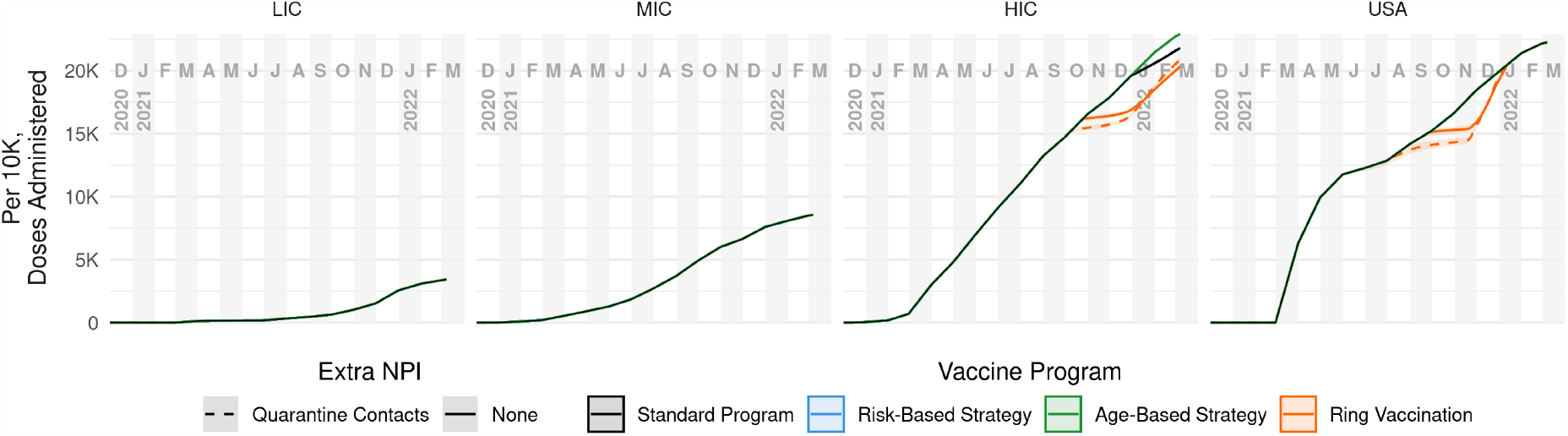
Cumulative vaccine doses administered per 10k over time, by supply level and distribution strategy. For each combination of the four supply levels LIC, MIC, HIC and USA (columns) and quarantine policy (dashed lines), we considered four vaccination strategies: ring vaccination (*i.e*., infection-risk prioritization) (orange), risk prioritization (blue), age prioritization (green), and a standard mass vaccination (black). For LIC and MIC levels, all strategies use all available doses. For HIC and USA levels, the strategies sometimes differ in doses delivered due to shortages of individuals eligible for revaccination; only risk- and age-based strategies always use all available doses.

Both quarantine and ring vaccination in the model rely on contact tracing. The contact tracing is intentionally imperfect: a majority (but not all) of reported cases are traced, and the number of contacts that are identified is dependent on the setting for that contact, *e.g*., all household members are identified, but a Poisson-distributed number of inter-household contacts will be identified. This process is repeated again for all of the identified contacts of the index case, thus both contacts and contacts-of-contacts may be identified; see S1 Additional Methods, Section 5.2 for details. We make simplifying assumptions that contact tracing capacity is unlimited, and that it can be completed the same day an index infection is detected, which could be days or weeks after infection occurred if *e.g*. detection occurs upon hospitalization or death. Real-world contact tracing will vary substantially between settings in capacity, speed, and thoroughness, but our assumptions offer a plausible upper bound on the effectiveness of quarantine and ring vaccination strategies.

A vaccination strategy can be thought of as a way to assign individuals to a queue to receive available vaccine doses. In the (passive) standard program, individuals are assigned at random to the queue. In the ring vaccination strategy, individuals are prioritized for vaccination if they are a contact-traced primary or secondary contact of an identified case. In the risk-based vaccination strategy, the position in the queue to receive vaccine is based on risk of severe disease, calculated using comorbid status and age, and quantized by decile. A cruder but more practical strategy that has often been used for COVID-19 is to assign risk based on age alone. We consider all combinations of vaccine supply and vaccination strategy factors with and without quarantine of identified cases, primary contacts, and secondary contacts. In S2 Additional Results, we also report the dynamics of a no-vaccine scenario, but do not use those for reference comparison in the main text results, as COVID-19 policy challenges have generally focused on how to use the doses available, and not whether to use them.

In all cases, we assume a three-dose vaccine regimen, with an mRNA-vaccine-like efficacy profile. This approximation is a simplifying assumption to reduce the complexity of the model and translation of empirical vaccine dose data into a model input. While many countries initially used nominally single dose vaccines, those products are generally lower efficacy (*17*) and many were ultimately deployed with additional booster doses, making their efficacy and dose requirements more like the mRNA products (see, *e.g*., (*18*)).

See S1 Additional Methods for details on determining the dose availability and distribution time series, and S2 Additional Results for simulated distribution stratified by dose ordinality and vaccinee age group.

To compare scenarios, we simulate *N* = 1000 replicates for each scenario, with random number generator seeds matched across scenarios. This ensures identical pre-vaccination-era histories when comparing across different scenarios. We compare matched replicates by calculating cumulative outcomes (infections and deaths) by incidence, incidence averted (compared to the reference program), and relative incidence averted (*i.e*., effectiveness) to date, then compute quantiles on these values. In S2 Additional Results, we provide similar measures for additional scenarios.

### Vaccine Efficacies and Dosing

Rather than simulate any particular SARS-CoV-2 vaccine product, we use published efficacy data (*17*) for different SARS-CoV-2 mRNA vaccines to approximate a generalized mRNA vaccine. We represent effects of vaccination using the efficacy parameters in Table S5 in S1 Additional Methods, namely VE_*S*_ (efficacy against infection), VE_*P*_ (efficacy against disease given infection), VE_*H*_ (efficacy against hospitalization given disease), and VE_*I*_ (efficacy against onward transmission given infection).

Note that the typical parameter reported from clinical trials is the unconditioned efficacy against disease, VE_*SP*_, which is related to VE_*S*_ and VE_*P*_ by Equation 1.

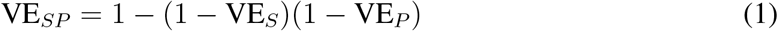

Each vaccination strategy determines how people become eligible for a first dose. On each day during a vaccination campaign vaccinees will be randomly drawn from the pool of all eligible people for each dose in the vaccination series. Vaccinees become eligible for second doses 21 days after the first dose, and eligible for third doses after a further 240 days (based on the median lag between second and third dose administration in Florida). Vaccinations on a given day continue until either no more doses are available, or no one is eligible.

## Results

### Model Calibration

We calibrated the model through an iterative process of algorithmic fitting and manual parameters adjustment. See Section 5.6 in S1 Additional Methods for details.

For a Florida-like population, we obtain reasonable matches to a range of metrics: observed values are mostly fluctuating about the central model trajectory and within the range suggested by the replicate trajectories (Fig 3). We use age- and dose-stratified vaccination data specific to Florida, USA for model calibration.

### Scenario Analysis

We used a detailed agent-based model to compare across different scenarios, considering effects of vaccine supply levels, vaccination strategies, and quarantine policy. Epidemic curves showing simulated outcomes over time illustrate gross features such as waves due to variants that emerge at specific time-points—alpha (*α*), delta (*δ*) and then omicron (*o*)—and also more subtle effects that arise from heterogeneities such as age-structuring with respect to comorbidities, employment-based interactions, and risk of death.

The simulations begin with the approximate start of the pandemic in most countries (early 2020); as we focus on the performance of vaccination campaigns, however, we show results only for the vaccination era in the model, December 2021 to March 2022. All scenarios give identical outcomes prior to the start of interventions. Fig 5 shows cumulative infections and deaths per 10K people during the vaccination era for four supply levels. Results for severe and critical disease have the same trends as those for deaths and are not shown. Fig 6 shows the same data in terms of overall cumulative effectiveness, computed relative to the performance of the standard (mass vaccination) program without quarantining. Effectiveness values of 0.1 (−0.1) indicate 10% fewer (more) adverse outcomes relative to the adverse outcomes under the standard vaccine program. The former figure indicates how the strategies perform in absolute terms, while the latter shows how much better (or worse) the more complicated strategies are than vaccinating people randomly.

**Figure 5:**
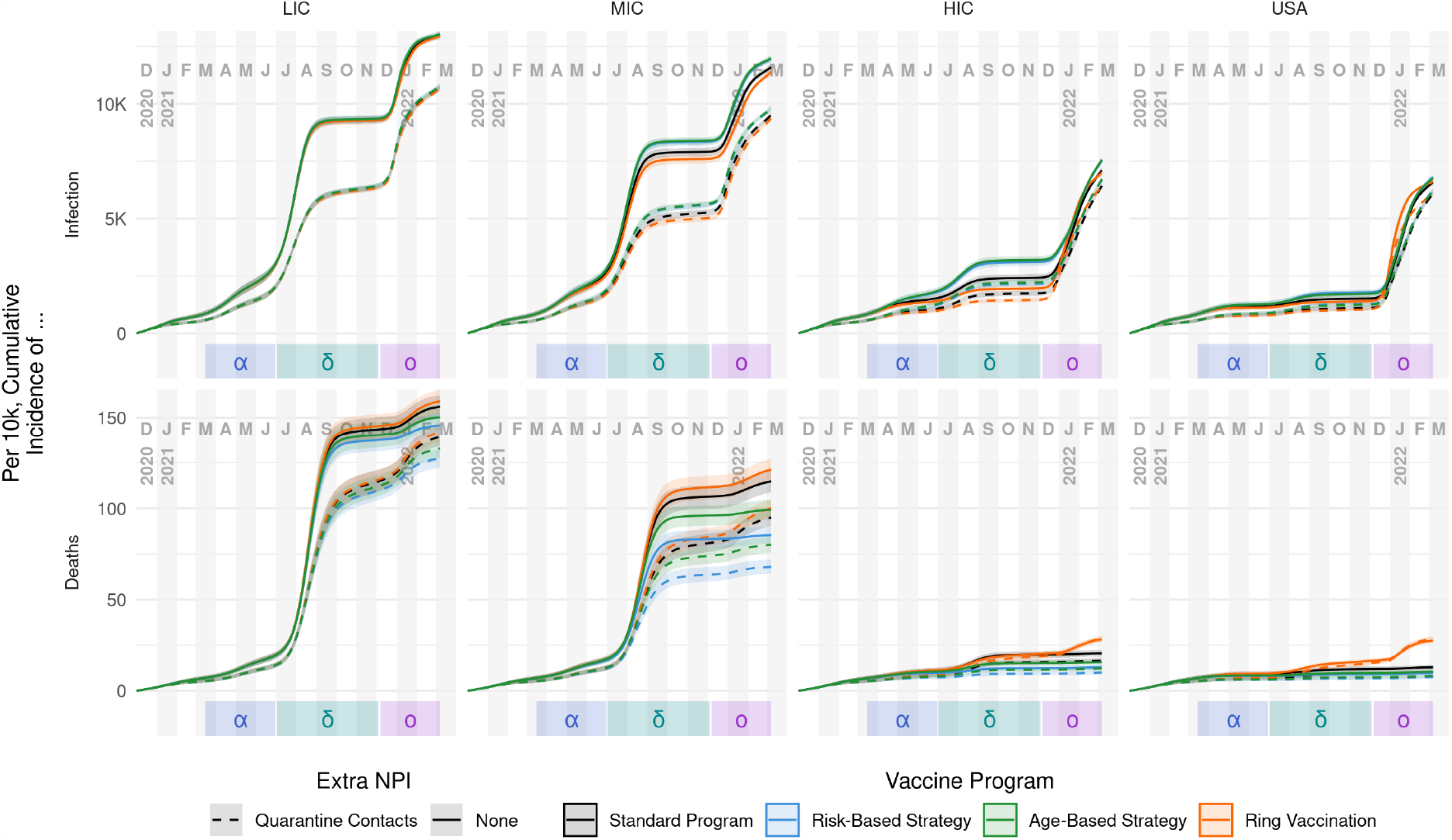
Cumulative incidence of infection and death per 10k people, by supply level and distribution strategy. Columns represent vaccine supply scenarios. Rows represent infection (top) and death (bottom) outcomes. Central lines represent median values with a 90% interquantile range shown as the ribbon. For infections, the major effects are supply level (columns) and the policy of quarantining (dashed lines) or not quarantining (solid lines), whereas the four vaccination strategies perform similarly. For deaths, supply level and quarantine are again the strongest factors. However, a strong effect of vaccination strategy also emerges: relative to a standard vaccine roll-out (black), risk-based vaccination (blue) and age-based vaccination (green) are more effective at preventing deaths, whereas ring vaccination (orange) is less effective. See the text for further explanation.

**Figure 6:**
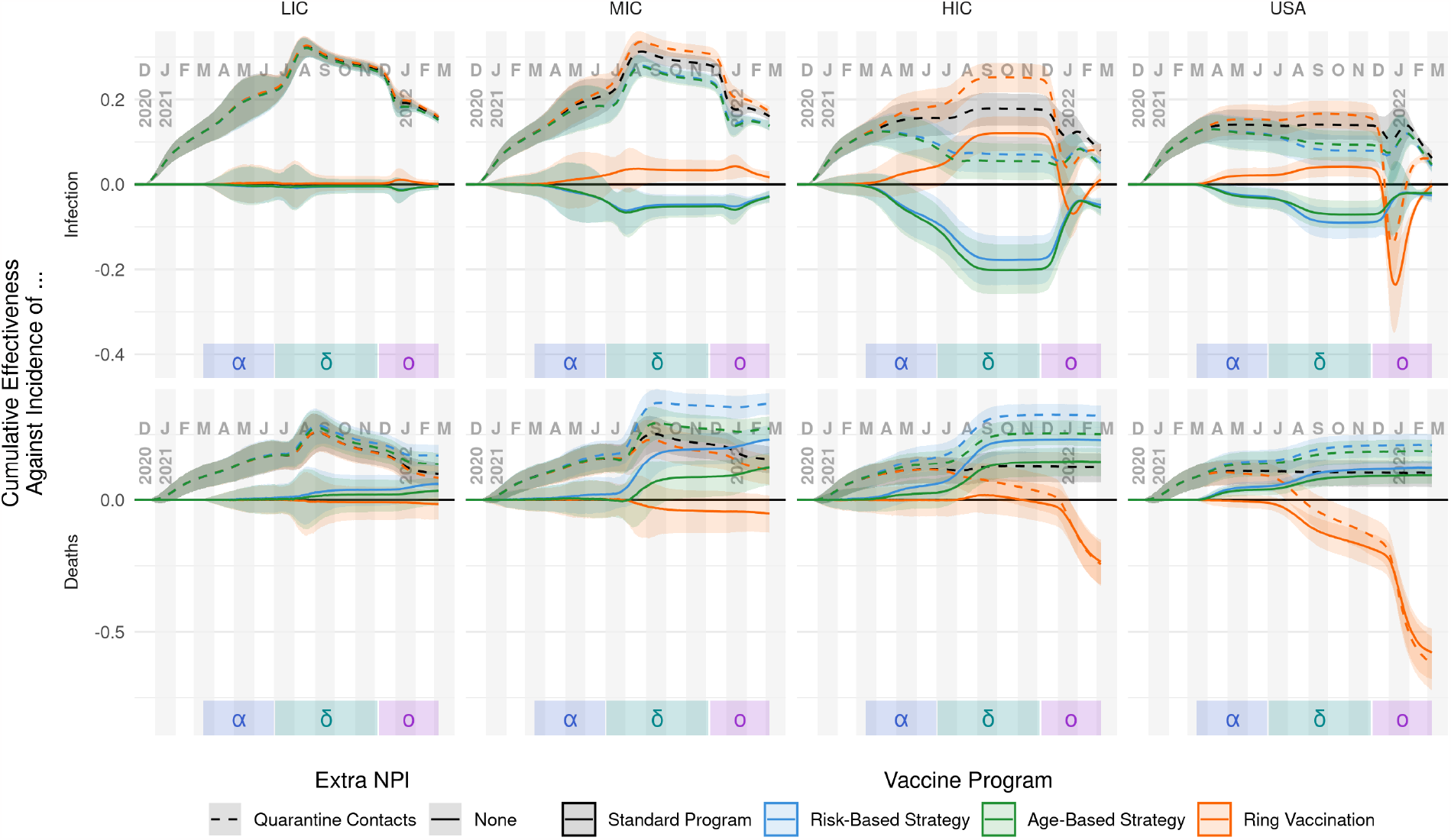
Cumulative overall effectiveness against infection and death incidence, by supply level and strategy. Columns represent vaccine supply scenarios. Against infections (top row), quarantining (dashed lines) significantly increases vaccination effectiveness. Central lines represent median values with a 90% interquantile range shown as the ribbon. Choice of strategy is less important in LIC and MIC scenarios, though in higher-income scenarios ring vaccination (orange) performs best until the omicron wave. Similarly, against deaths (bottom row), quarantining increases vaccination effectiveness overall; however, vaccination strategies are ranked more consistently. Risk- (blue) and age-based (green) strategies out-perform standard vaccination (black), while ring vaccination performs worst, especially in high-income settings during the omicron wave.

In absolute terms (Fig 5), the most important factor is vaccine supply, with increasing supply leading to lower absolute infections and deaths. The trends in relative terms are more complex: in lower supply settings, there are generally more remaining outcomes to be averted compared to the baseline, so large relative gains can be made with more sophisticated strategies. Essentially, each dose if used appropriately can have a proportionally larger effect, because there is more to be prevented. However, there are fewer doses available to make those gains, so the absolute impact is modest. This leads to clearer separation in relative performance, compared to the absolute perspective, and critically makes it easier to see strategy impact differences between cumulative infections and deaths.

For infections, all of the scenarios begin similarly, but incidence diverges rapidly in 2021. In high supply settings, overall coverage is sufficient to blunt infections during delta wave. Immunological resistance to omicron infection, to the extent that it existed, tended to come from delta infections in LIC and MIC settings, and from vaccination in HIC and USA settings (top and middle rows of Fig S7 in S2 Additional Results, respectively). Despite the significant discrepancies in vaccine supply and epidemic curves, the mean risk of infection given exposure was similar across all vaccine supply levels (bottom row of Fig S7 in S2 Additional Results) for the duration of the simulation. Because the model retains the complete infection and vaccination history for each person, it is possible to characterize the strain-specific extent and source of immunity throughout the simulated pandemic.

Quarantining of contact-traced individuals tends to dampen infections in the delta wave and earlier. Quarantine targets individuals most likely to be exposed (*e.g*., workers), however, who tend to differ from those most likely to suffer severe symptoms (*e.g*., seniors). When the omicron wave arrives, quarantining is not able to keep up with the increased transmissibility. Quarantine scenarios at that point have an immunity deficit compared to their no-quarantine counterparts, leading to a larger omicron wave.

However, the corresponding trends in deaths do not necessarily track with infections, as illustrated by comparing the top and bottom rows within Fig 5 and Fig 6. In general, the programs that somehow prioritize disease-risk (*i.e*., fully-risk-aware and age-based strategies) generally prevent fewer overall infections than even random distribution of the vaccine. Yet, they are also the most effective at preventing severe disease outcomes including death. Quarantining provided a consistent, if modest reduction in incidence of death—on the order of 10 to 15%— across supply levels and most vaccine programs, with the exception of ring vaccination in HIC and USA simulations, where quarantine had negative cumulative effectiveness after omicron. Apparent performance of a strategy depends on the timing of the assessment. We specifically considered comparison during periods of low transmission after epidemic waves as likely points of policy assessment. Fig 7 highlights trends in the simulation results, measured after each VOC wave. Generally, quarantining increases effectiveness regardless of vaccine supply, vaccination strategy, or VOC. Against infections, ring vaccination outperforms other vaccination strategies, though the effect is sometimes small and depends on both vaccine supply and assessment timing. Against deaths, a consistent vaccination strategy ranking (from best to worst performance) emerges regardless of vaccine supply or VOC: risk-based, age-based, standard, and ring vaccination. The discrepancy between most and least effective distribution strategy tends to increase with vaccine supply. Finally, for both infections and deaths, with and without quarantine and across all supply levels, the cumulative effectiveness of ring vaccination decreased as a result of the omicron wave. This is likely due to both the VOC spreading more rapidly (via increased infectiousness and decreased latent period), and decreased efficiency of the distribution strategy, as a smaller and smaller fraction of those traced would be as-yet unvaccinated and thus eligible for vaccination.

**Figure 7:**
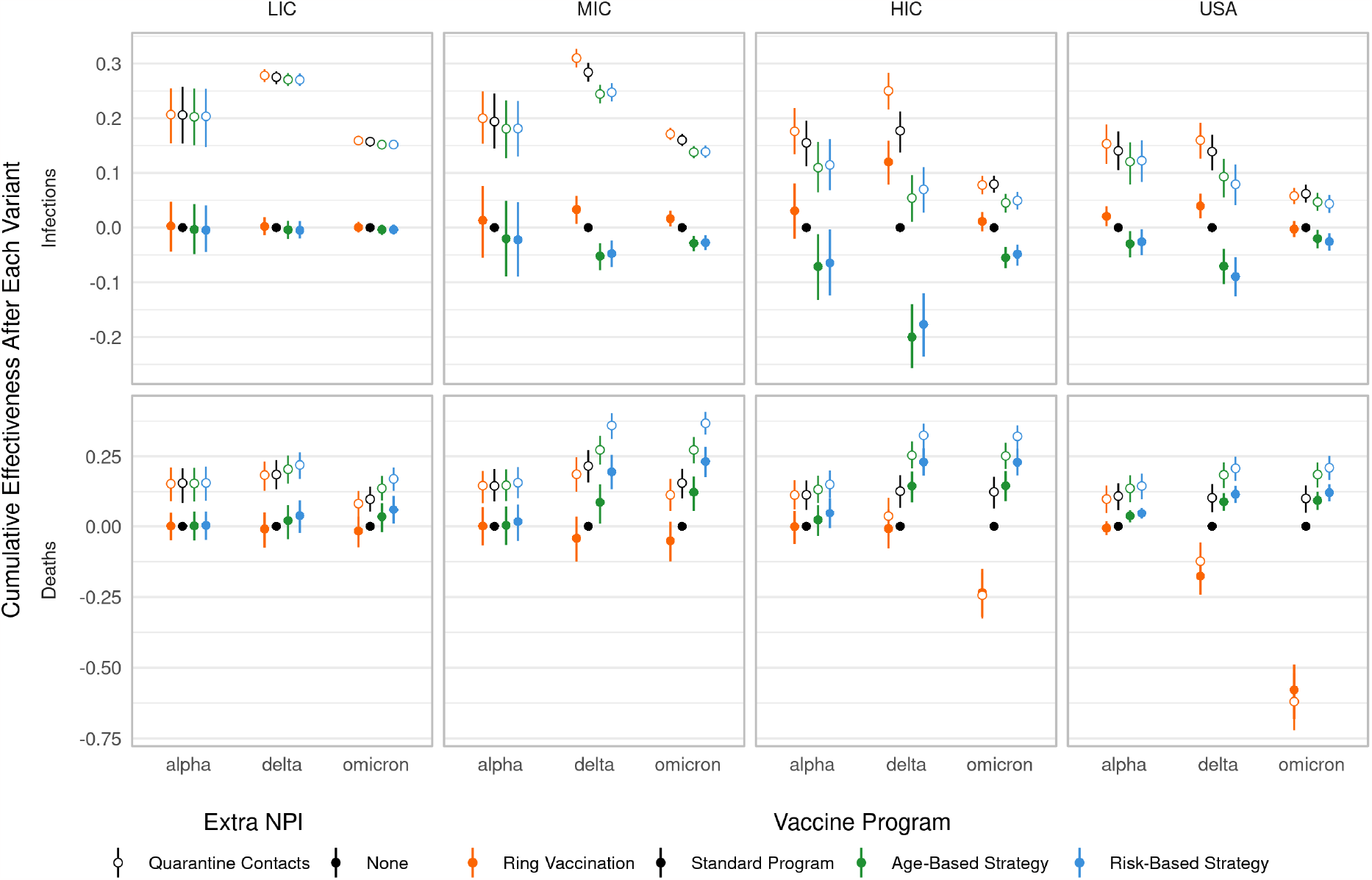
Cumulative effectiveness after variant waves. Columns depict vaccine supply scenarios and rows separate infection and death results. “Waves” are defined generally as the time from when a VOC is introduced to when a new VOC is introduced (however the alpha period starts at the beginning of the simulation and omicron period ends at the end of the simulation). The non-quarantining, standard strategy is used as the baseline for all comparisons.

Conditional vaccination, where vaccines are only given to individuals with no known infection history, provided modest additional benefits (compared to the main text, unconditional vaccination results) against death for LIC and MIC supply levels, particularly for risk-based vaccination (Figs S8 through S10 in S2 Additional Results).

## Discussion

Using a spatially-explicit, stochastic agent-based model, we were able to calibrate transmission parameters to reasonably match a wide-variety of outcomes (including reported cases, hospitalizations, deaths, seroprevalence, and breakthrough infections) for the COVID-19 pandemic in the state of Florida from February 2020 to March 2022. Using that calibrated model and population, we evaluated vaccination strategies that cover both the actual COVID-19 response programs and several alternatives. We evaluated overall strategy performance, *i.e*., at the scale of the entire population including non-vaccinees, in terms of infections and deaths. Strategies were compared to a “null strategy” standard program, where doses are administered randomly among the entire eligible population. We found consistent benefit from risk-prioritization programs for severe disease outcomes (*i.e*., disease warranting in-patient medical care and up to and including death), across supply levels, with and without additional quarantining NPIs. Strategy performance against infections was more nuanced, with program ranking depending on both socioeconomic setting and the timing of the assessment.

When only considering overall infections and not deaths, the risk-based programs tended to rank lower in cumulative effectiveness than alternatives during the alpha and delta SARS-CoV-2 waves, but the resulting increased immunity from those infections led to reduced omicron transmission, minimizing the differences between strategies by the end of the omicron wave (see Fig 6 and Fig S7 in S2 Additional Results). On the other hand, ring vaccination strategies, which specifically target segments of the population where transmission is occurring, demonstrated the reverse pattern, preventing infections due to earlier variants but losing ground against omicron. Notably, while ring-vaccination generally prevented the most infections, it under-performed even the standard program on severe outcomes, for all supply levels, unless supplemented with quarantining (see Fig 7). We expect that real-world use of ring vaccination for COVID-19 would be even less effective than our model’s predictions, particularly during peaks in transmission, as we did not assume resource limitations when contact tracing, nor delays in vaccinating traced individuals. Realistically, contact tracing capacity is specific to both locale and methodology, and whether ring vaccination would be practicable during *e.g*. an omicron-like wave would need to be evaluated given those specifics.

Increasing vaccine supply from the LIC to MIC to HIC average levels provided increasing benefit to all of the distribution strategies. Increasing vaccine supply from LIC to MIC levels reduced cumulative deaths by approximately 32% regardless of quarantining or vaccination strategy, and further increasing supply to HIC levels approximately reduced deaths by an additional 49%. However, these values assume an HIC-like population and infection-fatality rate (*19*); the absolute number averted would change with more context-specific assumptions, but the overall impact on relative changes is not obvious.

Against cumulative infections, the impact was somewhat less dramatic, largely because of the vaccine’s modest efficacy against infection and infectiousness. Increasing supply from LIC to MIC levels reduced cumulative infections by only 9%, while increasing to HIC levels reduced infections by an additional 25%. HIC and USA scenarios had similar cumulative vaccine coverage, and provided similar benefit for these relative reductions in infections and deaths.

For a given vaccine distribution strategy, adding quarantining as an NPI provided only modest additional benefit at HIC and USA vaccine supply levels (*<*10% reduction in infections and deaths). For lower supply levels, the impact was somewhat higher (approximately 10 to 15%). These are cumulative endpoints, measured after the omicron BA.1 wave. During the delta wave, we generally observed a bigger impact for quarantine, but quarantine was less effective against the more transmissible omicron variant. At the population scale, some of the infections that were avoided during delta were simply put off until omicron (see Fig 5 in S2 Additional Results). While the benefits of quarantine suggest a potentially attractive NPI strategy, that decision would need to account for costs (which might be worse in settings with lesser access to *e.g*., work-from-home alternatives) and transient threshold effects like running out of ICU beds, which we do not consider in this analysis.

Our approach has limitations, particularly when considering extrapolation to LIC and MIC settings. We fit model parameters to data from a particular HIC setting that (in comparison to typical LIC and MIC settings) is likely older and differs in comorbidities, economic and schooling activity patterns, and access to healthcare, among other distinctions. Using a detailed agent-based model makes it possible to evaluate equally detailed scenarios, but tailoring to specific settings requires both a large, varied collection of high-resolution non-epidemiological empirical data to construct the population (*e.g*., household structure, business distribution, schools structure) as well as detailed epidemiological time series. Our quantitative results for incidence of infections and deaths should not be interpreted as estimates for the LIC and MIC settings, as we do not have the relevant empirical data to calibrate the model. The relative change, *i.e*., effectiveness, should be more reliable, as prediction errors will tend to have similar direction and magnitude across scenarios for a given setting, and the ordered ranking of approaches should be most reliable.

Additionally, our model of risk perception and personal protective behaviors is essentially accounting for model residuals. When comparing the trends in inferred perceived societal risk to epidemic trends in our reference population, we see perceived risk slightly lagging incidence dynamics, with gradually dampening reactions. This matches our intuitions about how populations actually reacted to pandemic waves, so we are comfortable with this approach and suspect that attempts to compare to relevant empirical data (*e.g*., risk surveys, changes in mobility patterns) would be consistent. However, we did not attempt to codify this trend into a generic, reactive behavior model. Instead, we simply use the risk curves from the fitting stage for the alternative strategy scenarios. Those scenarios result in different incidence, which in real populations would likely affect risk perception and thus behavior. To deal with this issue, we would need to establish an explicit reaction model, ideally accounting for distinctions across income settings when extrapolating beyond the reference population. How best to model reactive behaviors, and what data to inform them with, remains an important open question in infectious disease modeling.

Overall, our model indicates that disease-risk prioritizing strategies consistently generated greater public health benefit than mass or ring vaccination. This finding did not vary with distribution setting, the addition of quarantining, or by timing of the measurement. The most demanding scenario in terms of information on disease risk provided the best results, though generally the less information-intensive age-prioritization appears to provide a sufficient surrogate for disease-risk. The actual best policy choice would incorporate other factors, for example vaccine distribution cost and political feasibility, which while not incorporated in this analysis, we would expect to favor simpler programs.

## Supporting information

SI 2 Additional Results

## Data Availability

All data produced in the present study are available upon reasonable request to the authors.
All code used in the present study is available online at http://github.com/tjhladish/covid-abm and https://github.com/kokbent/synthpop-fl

## Acknowledgments

This work was supported in part by The Emerson Collective, The Ron Conway Family, and NIH/NIAID (R56AI148284). Initial development of the agent based model was supported by the UFII COVID-19 SEED Fund. CABP was supported in part by the International Decision Support Initiative, which is funded by the Bill and Melinda Gates Foundation (OPP1202541), and by the World Health Organization (2022/1236532).

The National Corporation Directory provided data on Florida businesses including their locations and NAICS categories.

The authors acknowledge University of Florida Research Computing for providing computational resources and support that have contributed to the research results reported in this publication.

The identification of any specific commercial products is for the purpose of specifying a protocol, and does not imply a recommendation or endorsement by the National Institute of Standards and Technology.

