## Supplementary material for "Evaluating targeted COVID-19 vaccination strategies with agent-based modeling": SI 1 Additional Methods

### Supplement 2: Additional Results

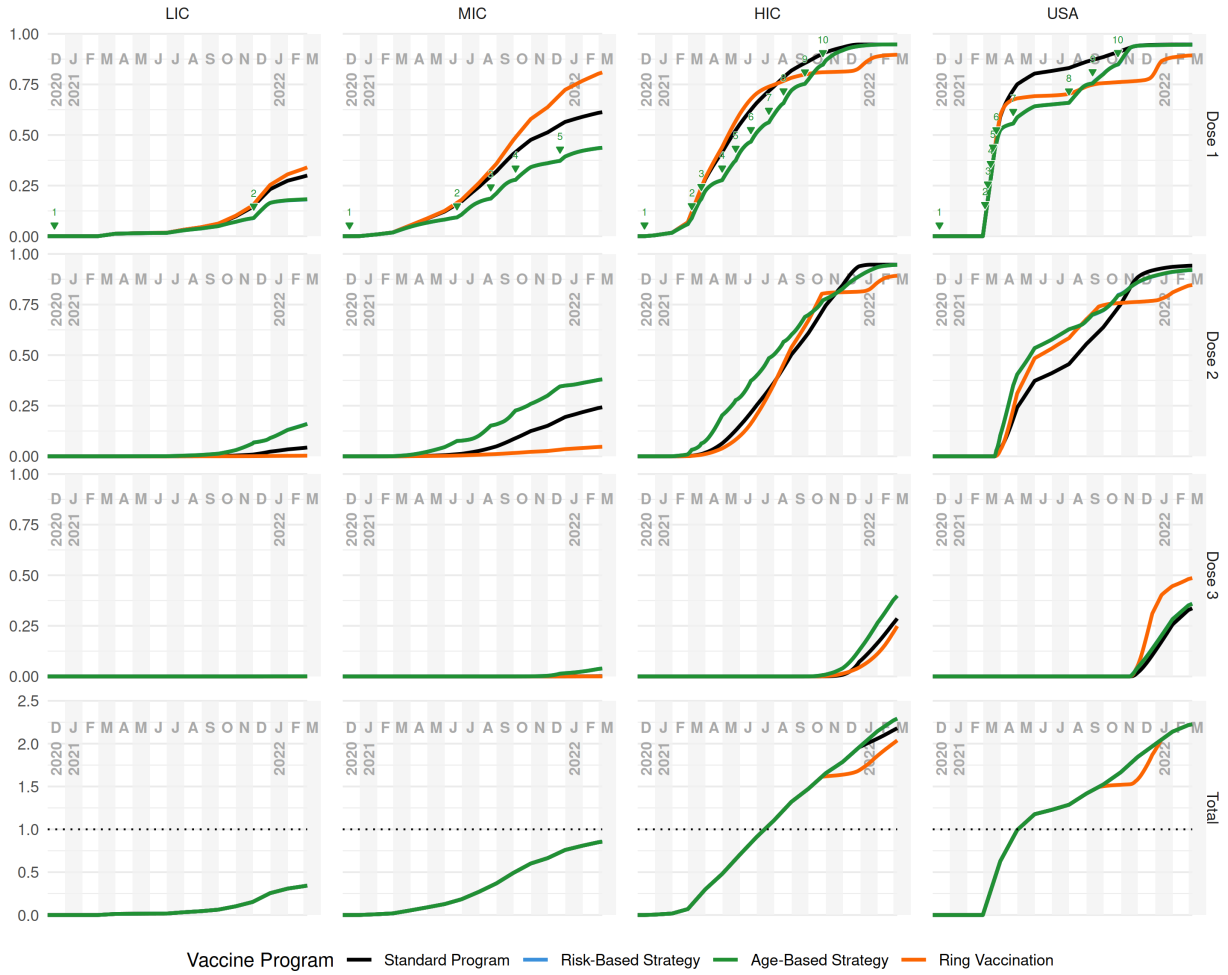


**Figure S1: Vaccine coverage by dose.** Columns represent vaccine supply scenarios and rows represent dose ordinality in the vaccine series (as well as total doses, row 4). Colors represent different vaccination strategies. In the Dose 1 row, the arrows depict the dates when age-decile groups are opened to be eligible for vaccination—the group addition dates are very similar for risk-based vaccination and so are not shown (see Section 6 in S1 Additional Methods for more details on vaccine strategies).

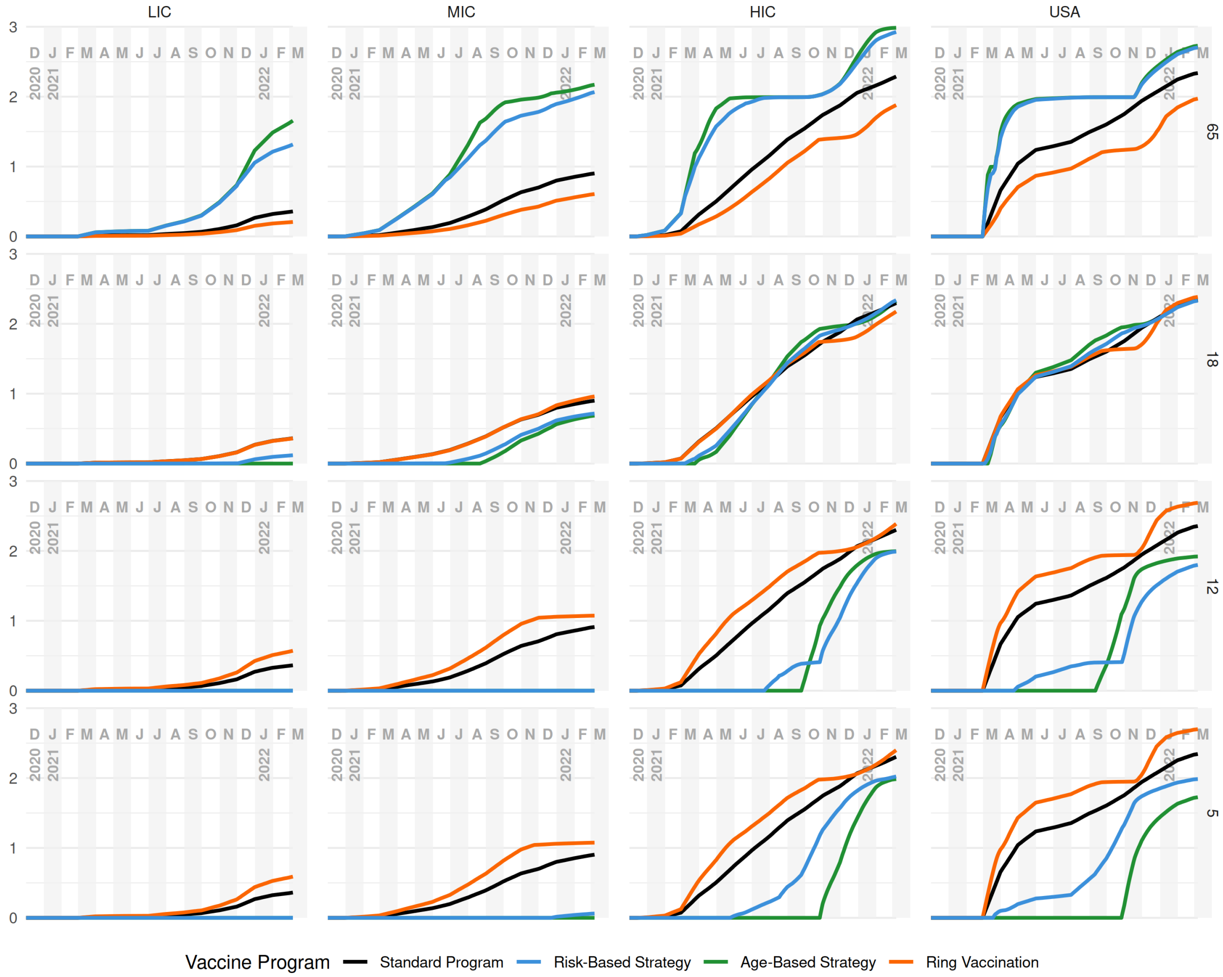


**Figure S2: Total vaccine coverage by age group.** Columns represent vaccine supply scenarios and rows represent age groups evaluated (top-down: 65+, 18–64. 12–17, 5–11). Colors represent vaccination strategies. Total coverage is calculated as the total number of doses cumulatively administered divided by the population of the age bin.

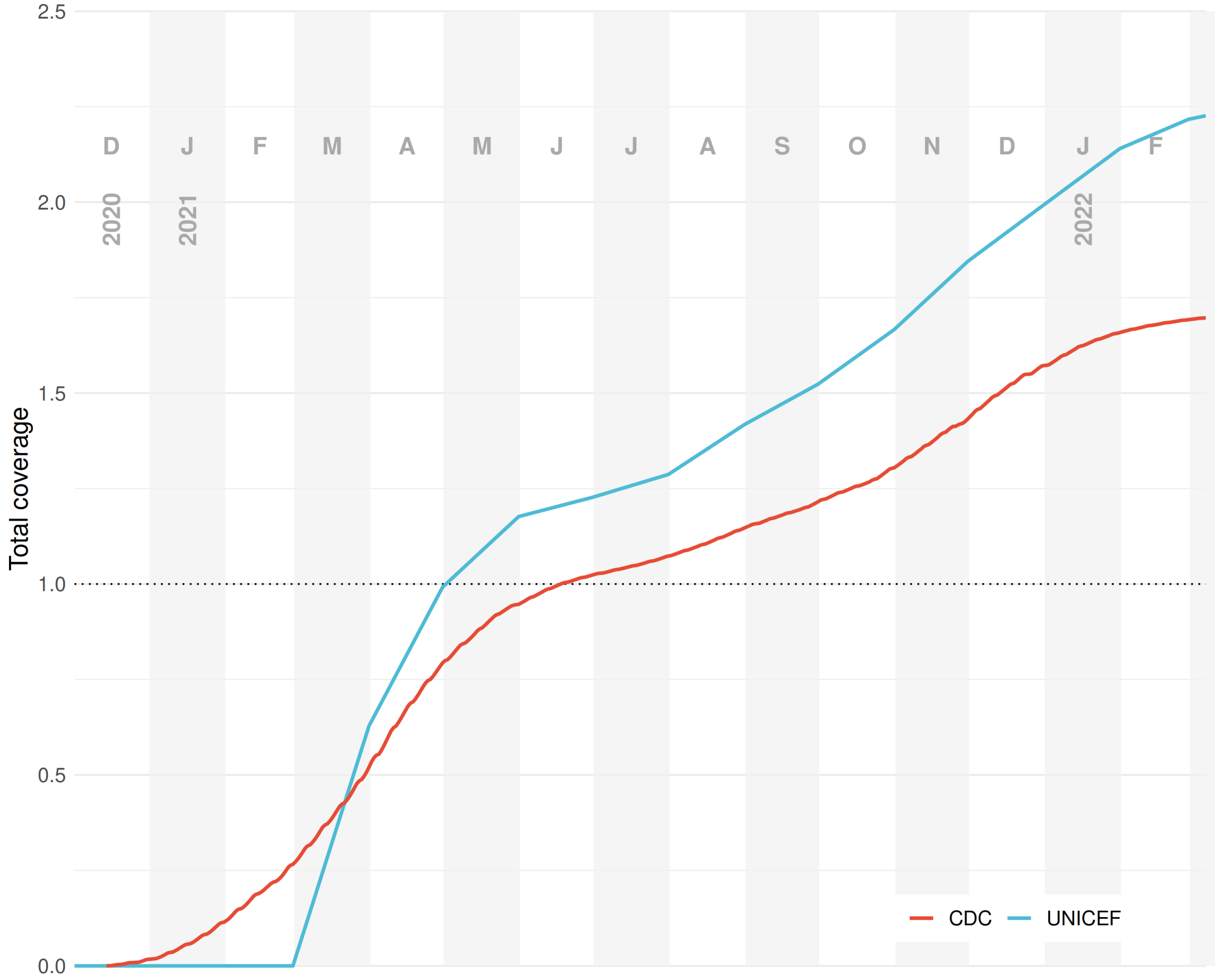


**Figure S3: UNICEF vs. CDC vaccine data comparison.** Both data sources represent total vaccine dose coverage (*i.e.*, total number of doses delivered) in the United States.

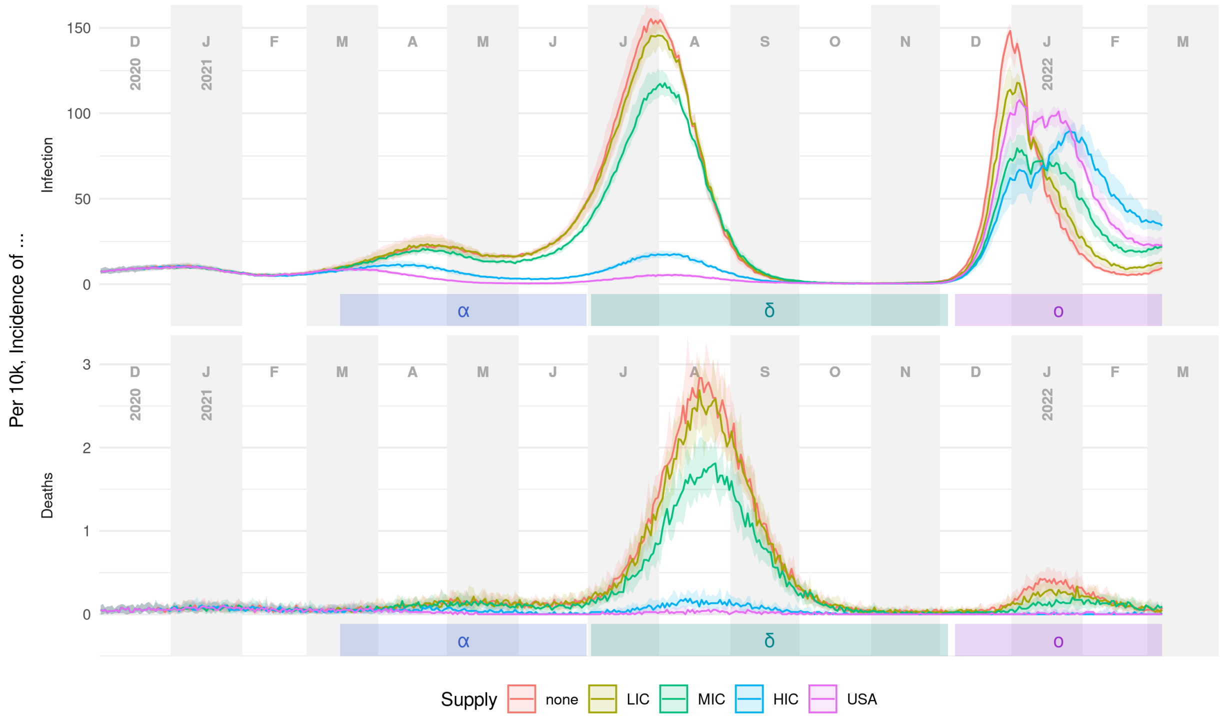


**Figure S4: Standard vaccine programs compared to no vaccination counterfactual.** A no-vaccination scenario (red) results in higher peaks in the delta and omicron waves. Standard programs here assume unconditional vaccination (as in the main text) without quarantining. Alpha, delta and omicron waves are noted using their Greek letters. Central lines represent median values with a 90% interquantile range shown as the ribbon.

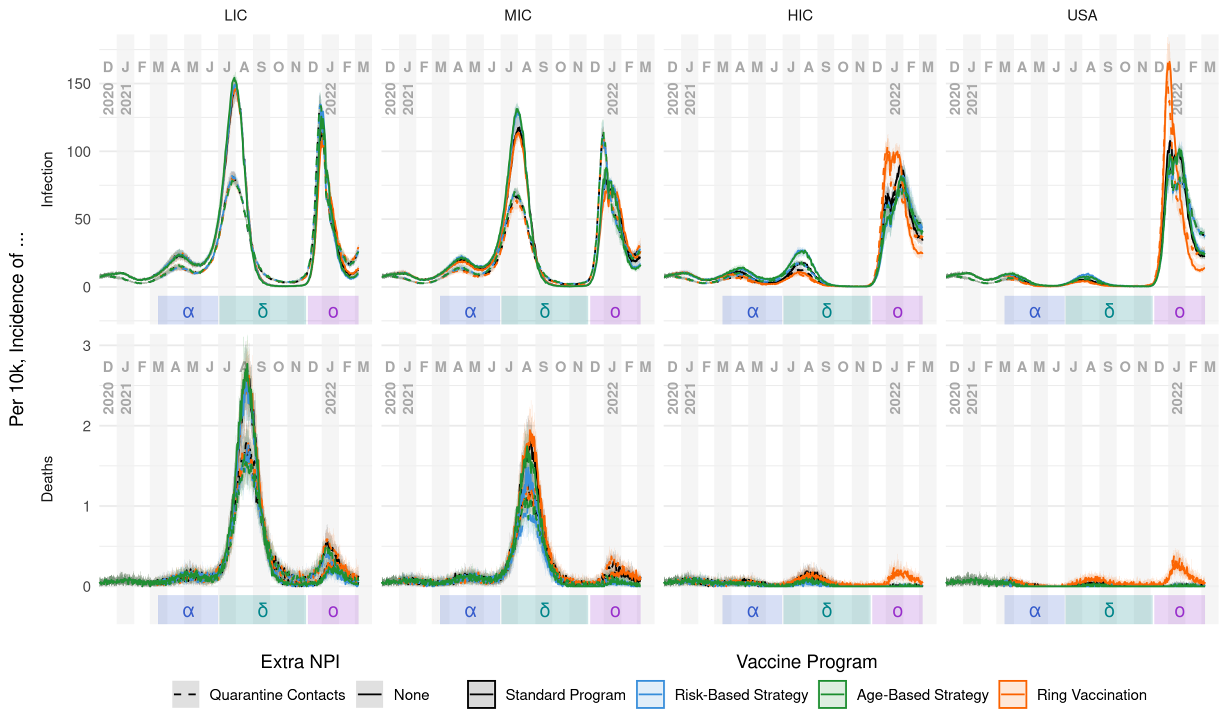


**Figure S5: Incidence of infection and death per 10k people, by supply level and distribution strategy using unconditional vaccination.** All data shown here use unconditional vaccination (*i.e.*, vaccinate any eligible people) in addition to other scenario features—similar to main text results. Central lines represent median values with a 90% interquantile range shown as the ribbon.

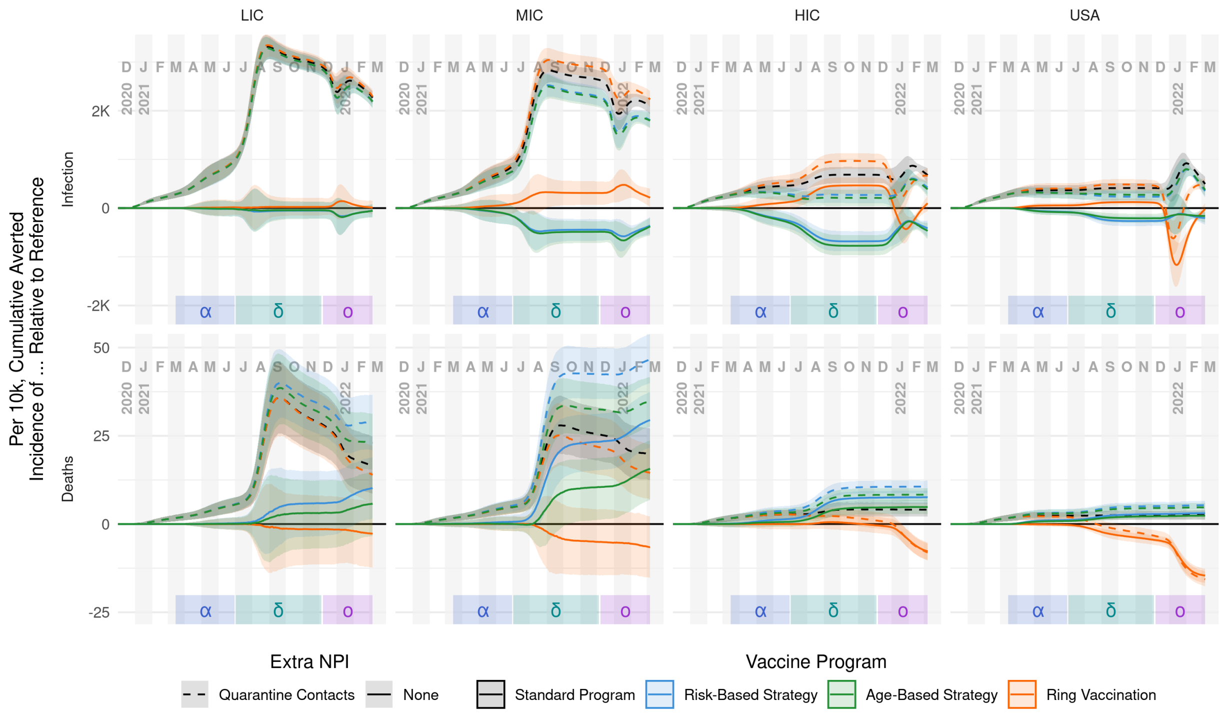


**Figure S6: Cumulative averted incidence per 10k people, by supply level and distribution strategy using unconditional vaccination.** This figure shows the difference (*i.e.*, outcomes averted) in performance between strategies, rather than the ratio (*i.e.*, effectiveness) that is reported in the main text. All data shown here use unconditional vaccination (*i.e.*, vaccinate any eligible people) in addition to other scenario features—similar to main text results. Central lines represent median values with a 90% interquantile range shown as the ribbon.

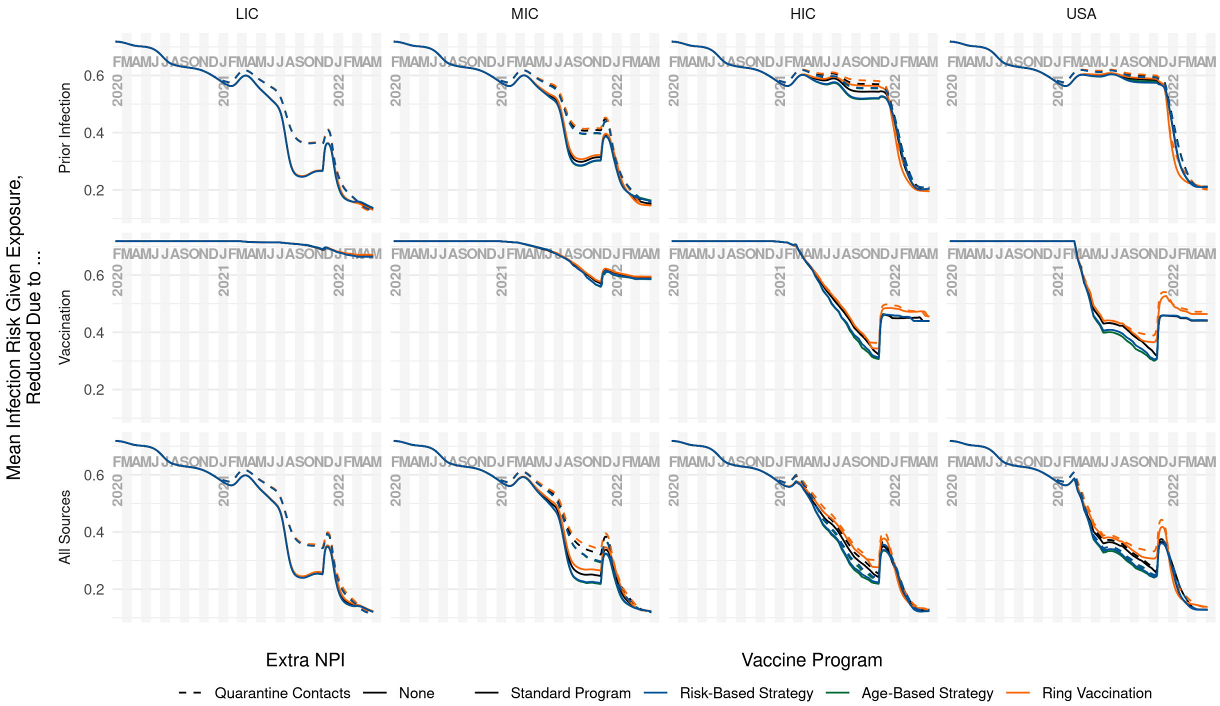


**Figure S7: Reduction in risk of infection due to immune protection.** Susceptibility to infection given exposure in the model (bottom row) is determined by age (not shown explicitly), infection history (top row), and vaccination history (middle row). Transient increases in risk are generally due to the emergence of immune escape variants, *i.e.* delta and omicron. Differences in risk across vaccine supply levels (bottom row) are modest, but the sources of immunity vary substantially: risk of infection in lower income settings was overwhelmingly due to having more infections in the past, whereas higher income settings acquired a similar overall level of risk reduction with more reliance coming from vaccinations, particularly during the delta wave. Within a single panel, differences between vaccine programs are due to the age structure of who has been vaccinated. Strategies that include quarantine generally result in higher risk because quarantine prevents some individuals from acquiring immunity via infection.

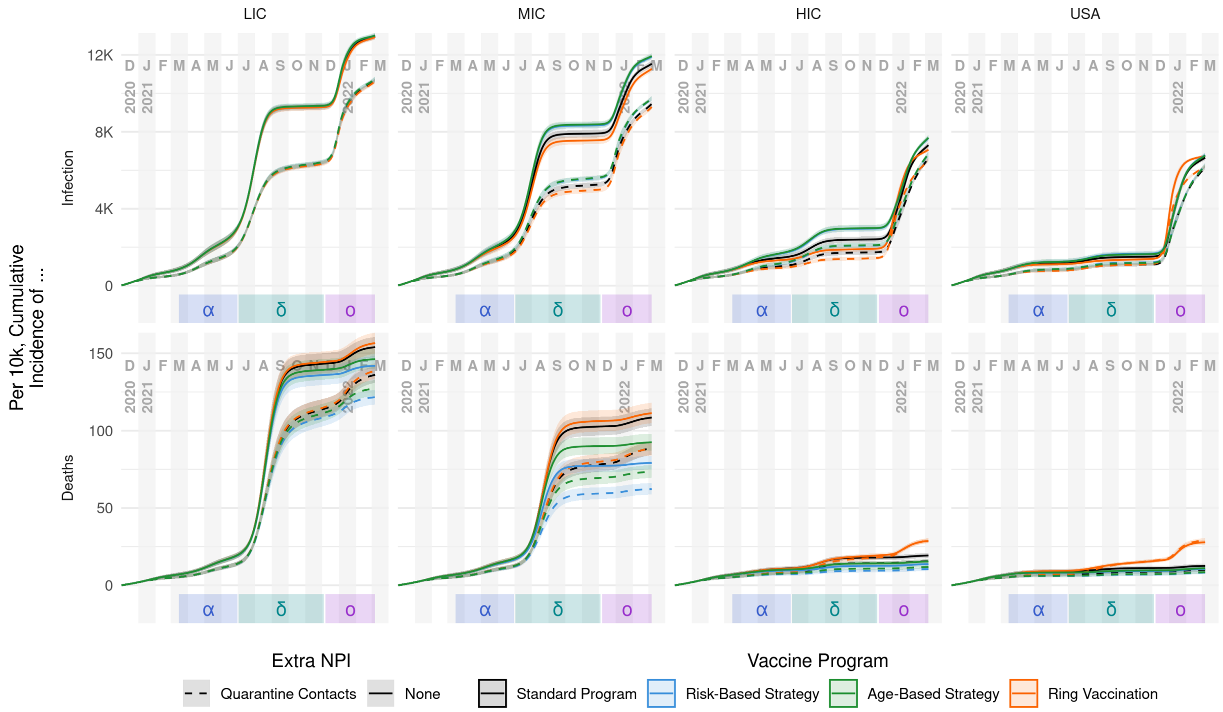


**Figure S8: Cumulative incidence of infection and death per 10k people, by supply level and distribution strategy using conditional vaccination.** All data shown here use conditional vaccination (*i.e.*, only vaccinate people with no prior case history) in addition to other scenario features. In regard to infections (upper row), the major effects are supply level (columns) and the policy of quarantining (dashed lines) or not quarantining (solid lines), whereas the four vaccination strategies perform similarly. In regard to cumulative deaths (lower row), supply level and quarantine are again the strongest factors. However, a strong effect of vaccination strategy also emerges: relative to a standard vaccine roll-out (black), risk-based vaccination (blue) and age-based vaccination (green) are more effective at preventing deaths, whereas ring vaccination (orange) is less effective. Central lines represent median values with a 90% interquantile range shown as the ribbon.

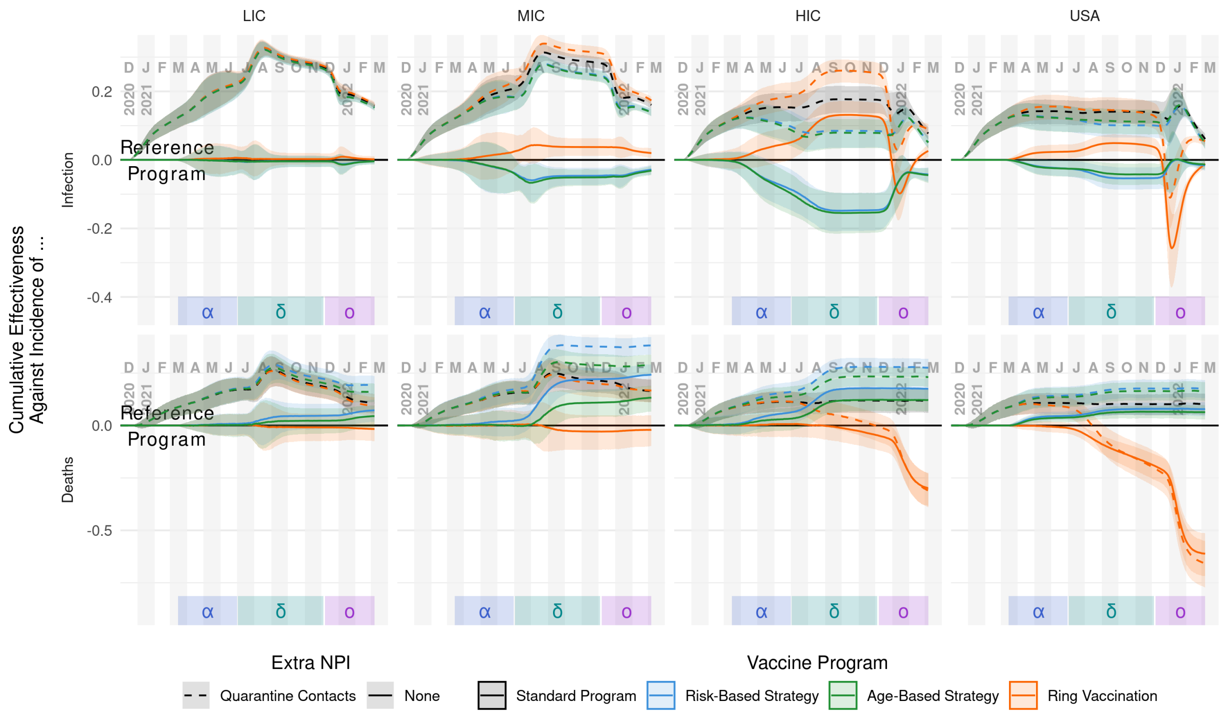


**Figure S9: Cumulative overall effectiveness against infection and death incidence, by supply level and distribution strategy using conditional vaccination.** All data shown here use conditional vaccination (*i.e.*, only vaccinate people with no prior case history) in addition to other scenario features. Against infections, quarantining (dashed lines) significantly increases vaccination effectiveness. Choice of strategy is less important in LIC and MIC scenarios, though in higher-income scenarios ring vaccination (orange) performs best until the omicron wave. Similarly, against deaths, quarantining increases vaccination effectiveness overall; however, vaccination strategies are ranked more consistently. Risk- (blue) and age-based (green) strategies out-perform standard vaccination (black), while ring vaccination performs worst (especially in high-income settings during the omicron wave). Central lines represent median values with a 90% interquantile range shown as the ribbon.

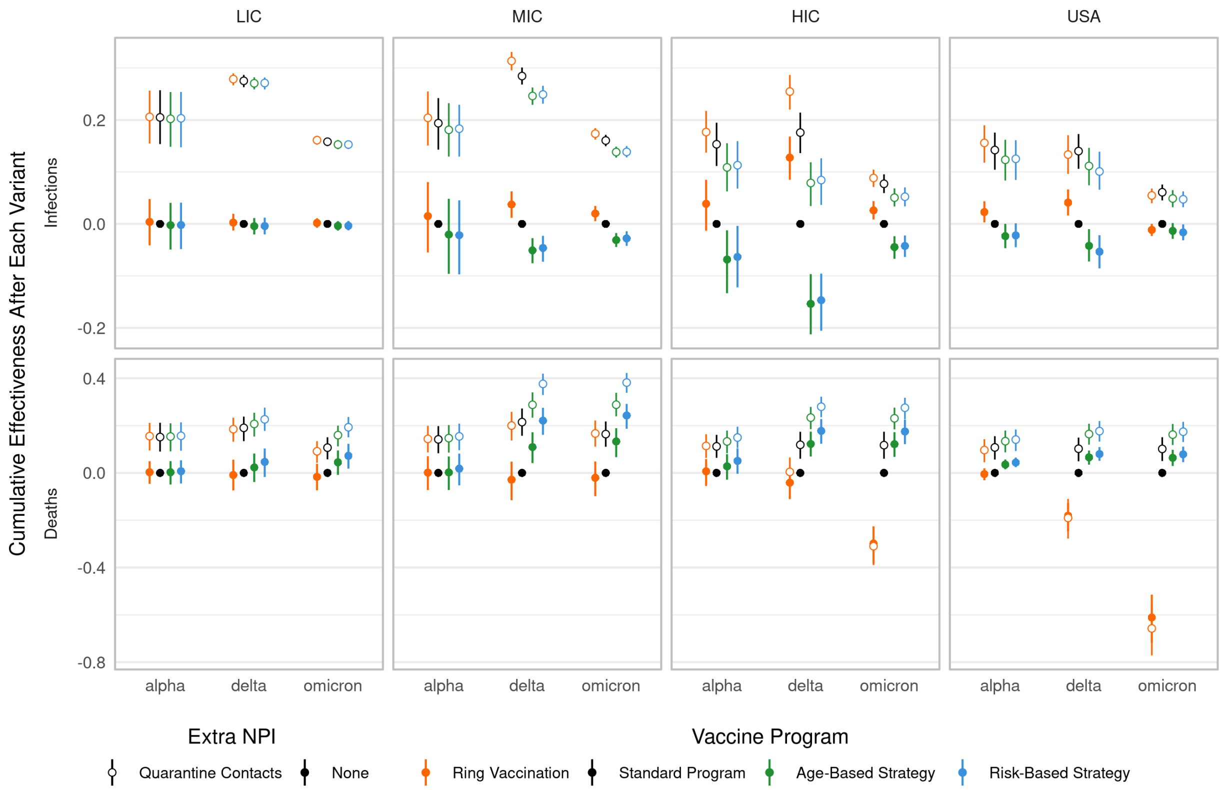


**Figure S10: Cumulative effectiveness after variant "waves" using conditional vaccination.** All data shown here use conditional vaccination (*i.e.*, only vaccinate people with no prior case history) in addition to other scenario features. "Waves" are defined generally as the time from when a VOC is introduced to when a new VOC is introduced (however the alpha period starts at the beginning of the simulation and omicron period ends at the end of the simulation). The non-quarantining, standard strategy is used as the baseline for all comparisons.
