## Supplementary material for "Evaluating targeted COVID-19 vaccination strategies with agent-based modeling": SI 2 Additional Results

### Supplement 1: COVID-ABM Description

#### 1 Model Overview

We extended an agent-based model framework to support evaluating the COVID-19 epidemic and response efforts. The model includes empirically-informed SARS-CoV-2 mechanisms in a stochastic, discrete-time transmission process in a population of agents moving between places. This model was derived from the model of (*1*); an early version was used by (*2*). The model has been developed to represent the entire state of Florida (*i.e.*, 20.6M people and 11.2M locations), but for generalizability and computational practicality in this analysis, we used a representative sub-population of 375K people, with spatial structure corresponding to Marion County, Florida. The 162K households in the model are thus sampled from the population of the entire state, with features including household size, ages in years, and employment or student status. The presence of comorbidities relevant to COVID-19 is known in aggregate but not at the household level, and thus is sampled independently. Location types in the model aside from residences are based on their actual addresses in Marion County, including 33.8K workplaces, 118 schools, 46 long-term care facilities, and 6 hospitals. We calibrate the outcomes of this population to match the state-wide dynamics.

People in the model have a range of behaviors and interactions that may result in transmission. Some interactions always occur, like those within households, hospitals, and long term care facilities. Interactions in schools and non-essential workplaces may be reduced via top-down non-pharmaceutical interventions (NPIs), while those due to social interactions and patronage of workplaces may be avoided via personal protective behaviors (PPBs). Infection outcomes in the model are affected by age and the presence or absence of comorbidities.

Time in the model is discrete, following a daily cycle of transmission opportunities based on the co-localization of infectious and susceptible individuals in households, workplaces (both as employees and patrons), schools, long-term care facilities, and hospitals. This approach leads to age- and locality-specific interaction patterns, *e.g.*, children attend specific schools during the week, where they interact with other children from the model's catchment area associated with that school. Social interactions in the model are also structured, as individuals with similar levels of COVID-19 risk aversion are likely to be "friends", defined as having specific inter-household opportunities for transmission independent of residence, work, school, and business patronage (Fig 2 in Main Text).

### 2 SARS-CoV-2 and COVID-19 Natural History

In our model, individuals begin in a partially susceptible state, with a level of leaky, innate immunity that is age-specific, *i.e.*, an individual may probabilistically resist infection from one exposure, but become infected via a different interaction later in the same day. Age-specific innate immunity is based on the inferred values in (*3*). Individuals who are exposed to an infected individual (with an introduced or locally-acquired infection) may become infected and proceed, via sequential stages, to an asymptomatic, mild, severe, or critical infection. Individuals who are asymptomatic or with mild or severe symptoms may recover directly, whereas those with critical infections must revert back to a severe infection before recovery. Death occurs only in critical cases (Fig 1a in Main Text). Severe cases may be hospitalized, and critical cases may be admitted to an intensive care unit (ICU; see Section 3.5). Recovered individuals have acquired, leaky immunity to further infection that is strain-specific and changes over time. Acquired immunity, whether derived from prior infection or vaccination, may reduce the probability of infection, onward transmission, and the probabilities of progressing to more severe states should infection occur (see Section 2.2).

We use existing literature to inform natural history parameters as much as possible, *e.g.*, disease state durations are sampled from distributions informed by meta-analyses and published models; see Table S1. Infection severity is probabilistic, depending on an individual's age and presence of relevant comorbidities, as summarized in Table S2.

| Transition | Duration distribution | Source |
|---|---|---|
| infection → symptoms | Gamma(3.1, 1.6) | (*4*) |
| infection → infectiousness | Binom(0.53, incubation_period - 1) + 1 | Fit to (*5*) |
| symptoms → clearance [a] | 7 (for wildtype) | (*5*) |
| infectiousness → clearance [b] | Calculated based on above values | Assumed |
| mild → recovery [c] | Gamma(4.1, 2.8) | Fit to (*6*) |
| mild → severe | Gamma(0.81, 7.2) | Fit to (*7*) |
| severe → recovery [d] | empirical CDF [e] | (*8, 9*) |
| critical → severe | empirical CDF [e] | (*8*) |
| critical → death | NBinom(0.2,2) | Fit to (*10*) |

**Table S1: State duration parameters.** Transitions between disease states specified in the first column are given by the distribution of state durations (in days) specified in the second column. Gamma distributions are specified using (shape, scale) parameters. Mild disease is here defined as clinically ill, but not warranting hospitalization. Severe and critical disease warrant hospitalization and intensive care, respectively, but receiving medical care is probabilistic. Neither hospital nor ICU admission affect disease-state durations, only the probability that a patient recovers rather than dies. [a]Clearance refers to the end of infectiousness, but not necessarily the end of symptoms, whereas recovery indicates end of symptoms. [b]The infectious period is calculated as the incubation period plus the symptomatic period minus the latent period. This calculation is always used; whether the infection actually becomes symptomatic is determined independently. [c]Used if symptoms do not become severe. [d]Severe state duration does not include critical state duration, and is estimated by subtracting the ICU stay duration distribution provided by (*9*) from the total hospital stay duration provided by (*8*). For individuals who become critically ill, we assume half the severe state is spent before becoming critically ill, and half afterward, assuming the individual does not die while critically ill. [e]CDF, cumulative distribution function.

In the US, individuals with an underlying health condition have six-fold higher chance of developing severe infection requiring hospitalization and 12-fold higher chance of death compared to individuals without any comorbidity (*11*). This list of underlying health conditions is extensive; the common comorbidities with sufficient empirical data to inform our model are cardiovascular disease, diabetes, chronic lung disease, and severe obesity (*11*). For the purposes of determining infection prognosis, we combine all comorbidities and assign each individual a comorbidity state of true or false. See Section 3.1 for details on how we determine the distribution of comorbidities.

| Conditional transitions | Probability | Source |
|---|---|---|
| Pr{infection \| exposure} | Varies by age | (*3*) |
| Pr{symptomatic illness \| infection} | Varies by age | (*3*) |
| Pr{severe illness \| symptomatic illness} | Varies by age & comorbid status | (*11, 12*) |
| Pr{critical illness \| severe illness} | Varies by age & comorbid status | (*11, 12*) |
| Pr{hospitalization \| severe illness, general pop.} | 0.8 | Assumed |
| Pr{hospitalization \| severe illness, LTCF pop.} | 0.25 | Assumed |
| Pr{ICU admission \| hospitalization, critical illness} | 0.9 | Assumed |
| Pr{ICU admission \| non-hospitalized, critical illness} | 0.75 | Assumed |
| Pr{death \| critical illness, non-ICU} | 0.9 (for wildtype) | Assumed |
| Pr{death \| critical illness, ICU} | Varies by age & comorbid status | (*11, 12*) |

**Table S2: Transition probability parameters.** Symptomatic illness is defined as disease that would typically be recognized by a clinician. Comorbid status is binary, based on the presence or absence of other disease that affects COVID-19 prognosis. LTCF: long term care facility; ICU: intensive care unit.

#### 2.1 SARS-CoV-2 Variants

Each infection in the model is caused by a specific SARS-CoV-2 variant, with variant type passed from infector to infectee for locally-acquired infections. New variants appear in the population via external introductions. We manually choose introduction start dates that result in strain displacement rates that are consistent with empirical data, *e.g.*, alpha reached approximately 50% prevalence in Florida by March 2021, delta by July 2021, and omicron (BA.1) by December 2021 (*13*). Initially, all infections are caused by a strain we refer to as the wildtype.

We address the appearance of new variants by adjusting the variants used in external introduction events, and do not model mutation within hosts. Specifically, external introduction events switch from one variant to the next (e.g., alpha to delta) over a specific calendar interval, in a linearly graduated manner. As all individuals have an equal, small probability ($p = 10^{-4}$) of exposure daily from an external source, the number of exposure events on a given day is binomially distributed. Variants are sampled for these exposures in a weighted, graduated manner, to represent the increasing external prevalence of new variants. For the switch from wild-type to alpha or alpha to delta, the duration of the transition is 50 days, with the probability of sampling the new variant increasing linearly by 2% each day. For the delta to omicron transition, the switch takes place over 10 days, *i.e.*, a 10% increment per day. The resulting distribution of variants introduced on a given day thus reduces to a single type for much of the pandemic, and is binomial for the VOC timings that we assume during transitions.

Variant prevalence in the model changes organically as a result of the changing introduction probabilities, differences in variant properties, and differences in population-level immunity to specific variants due to past infection or vaccination (see Section 2.2).

| Parameter | Value | Source |
|---|---|---|
| | wildtype | |
| Infectious hazard | 0.07 | Table S6 |
| Pathogenicity | varies by age | Table S2 |
| Immune escape probability | 0.0 | Assumed |
| Severity | varies by age & comorbid status | Table S2 |
| ICU mortality | varies by age & comorbid status | Table S2 |
| Incubation pd. | Sampled | Table S1 |
| | Alpha | |
| Relative infectiousness | 1.6 * wildtype | (*14*) |
| Relative pathogenicity | 1.1 * wildtype | (*14*) |
| Immune escape probability | 0.15 | (*14*) |
| Relative severity | wildtype | See above |
| Relative ICU mortality | wildtype | See above |
| Symptomatic infectious pd. | wildtype | See above |
| Relative symptom onset | wildtype | See above |
| | Delta | |
| Relative infectiousness | 1.6 * alpha | (*15*) |
| Relative pathogenicity | 2.83 * alpha | (*15*) |
| Immune escape probability | 0.2 | (*15*) |
| Relative severity | 1.4 * wildtype | (*15*) |
| Relative ICU mortality | 3.0 * wildtype | (*15*) |
| Symptomatic infectious pd. | wildtype | See above |
| Relative symptom onset | wildtype | See above |
| | Omicron | |
| Omicron/wildtype infectious pd. ratio | 2/3 | |
| Relative infectiousness adjustment | calculated | See Eqn S1 |
| Relative infectiousness | 2.0 * delta / relative_infectiousness_adjustment | (*16, 17*) |
| Relative pathogenicity | 0.5 * delta | (*16, 17*) |
| Immune escape probability | 0.5 | (*16, 17*) |
| Relative severity | 0.5 * delta | (*16, 17*) |
| Relative ICU mortality | 1.5 * wildtype | (*16, 17*) |
| Symptomatic infectious pd. | 8 days | |
| Relative symptom onset | 0.5 * wildtype | (*16, 17*) |

**Table S3: VOC parameters.** In general, values in this table were initially based on estimates from the citations provided, then hand-adjusted to match Florida outcomes. The relative adjustment for infectiousness of omicron is given by Equation S1.

Relative infectiousness of strains is typically expressed in terms of the entire infectious period. As we use a hazard model of transmission, and we assume omicron has a different (shorter) infectious period from previous variants, we need to increase omicron's transmission hazard so that has the overall relative infectiousness we want. This relative infectiousness adjustment is calculated according to Equation S1, where $\tau_H$ is the wildtype infectious hazard and $\kappa_v$ is the infectious period of strain $v$.

$$relative\_infectiousness\_adjustment = \frac{1.0 - (1.0 - \tau_H)^{\kappa_o/\kappa_{wt}}}{\tau_H} \tag{S1}$$

#### 2.2 Immunity

Both infection- and vaccine-derived immunity against future infection ($\text{IE}_S$ and $\text{VE}_S$, respectively) are modeled as a leaky process with short-term and long-term stages, in which prior immunity reduces the per-exposure probability of infection (Fig S1). Infection- and vaccine-derived immune protection against pathogenicity, severe outcomes, and onward transmission are modeled as static parameters (see Tables S4 and S5), although these values are modulated by strain-specific coefficients (Table S3).

| IE parameters | Value | Reduction in |
|---|---|---|
| $\text{IE}_S$ | Varies | susceptibility |
| $\text{IE}_P$ | 0.75 | Pr{pathology \| infection} |
| $\text{IE}_H$ | 0.5 | Pr{severe outcomes \| pathology} |
| $\text{IE}_I$ | 0.0 | infectiousness |

**Table S4: Infection efficacy (IE) parameters.** $\text{IE}_S$ is specific to infections history and challenging strain, and varies by individual; see Section 2.2.1 for details. Values for other types of efficacy are assumed.

##### 2.2.1 Infection-derived Immunity

We model prior infection-based immunity against future infection using three distinct filters: broad, short-term immunity; strain-specific, long-term immunity; and broad, long-term immunity (see Fig S1). If an individual has an infection history, upon every subsequent exposure, these filters are checked to determine whether one of them prevents the exposure from causing infection.

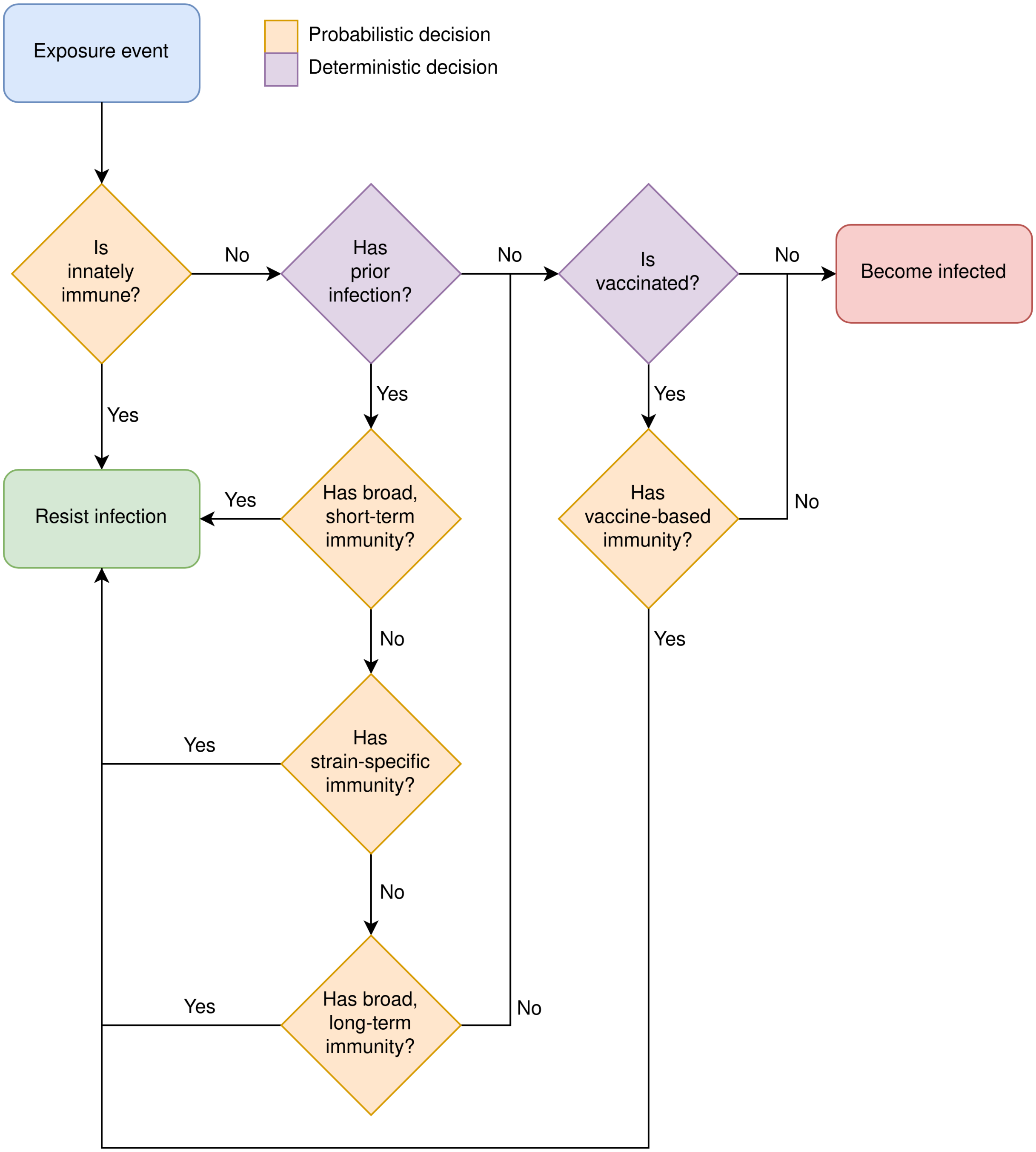


**Figure S1: Infection decision flowchart.**

**Broad, Short-term Immunity** Every individual is assigned a baseline neutralizing antibody titre, which is log-normally distributed (log-mean of 0, log-standard deviation of 1, then multiplied by 2; *i.e.*, $X \sim 2 \times \text{Exp}\left(\text{Norm}\left(0, 1\right)\right)$). That antibody titre is translated into an efficacy against infection based on empirical relationships between neutralizing titre and efficacy (*18*). Short-term immunity is modeled as a leaky process against an individual's starting infection protection. This comparison is further modified by the immune-escape probability of the viral strain involved in the exposure event (see Table S3).

**Strain-specific, Long-term Immunity** If the challenging strain matches that of a past infection, there is an additional chance for immune protection. The exposure is resisted if the time since the last matching infection is less than 60 days, or based on a leaky process with a probability of 0.85 if the interval is at least 60 days (based on findings in (*19*)).

**Broad, Long-term Immunity** Individuals vary in their inherent ability to develop broad spectrum, effective immunity after infection. Every individual is assigned a general immune cross-protection probability sampled from a Uniform(0.0, 1.0) distribution. If the individual's cross-protection probability is less than the immune-escape probability of the challenging viral strain (see Table S3), the exposure is resisted.

##### 2.2.2 Vaccine-derived Immunity

Vaccine-derived immune protection is modeled similarly to infection-derived immunity. Vaccine protection against infection is modeled as a leaky process involving initial vaccine efficacy against infection augmented by the immune-escape probability of the exposure's viral strain. This immune protection is only active 10 days after a vaccine dose is received.

#### 2.3 Vaccine Performance

We represent effects of vaccination using the efficacy parameters in Table S5, namely $\text{VE}_S$ (efficacy against infection), $\text{VE}_P$ (efficacy against disease given infection), $\text{VE}_H$ (efficacy against hospitalization given disease), and $\text{VE}_I$ (efficacy against onward transmission given infection).

Note that the typical parameter reported from clinical trials is the unconditioned efficacy against disease, $\text{VE}_{SP}$, which is related to $\text{VE}_S$ and $\text{VE}_P$ by Equation S2.

$$\text{VE}_{SP} = 1 - (1 - \text{VE}_S)(1 - \text{VE}_P) \tag{S2}$$

| Parameter | Dose 1 | Dose 2 | Dose 3 | Reduction in |
|---|---|---|---|---|
| $\text{VE}_S$ | 0.40 | 0.80 | 0.80 | susceptibility |
| $\text{VE}_P$ | 0.67 | 0.75 | 0.75 | Pr{pathology \| infection} |
| $\text{VE}_{SP}$ | 0.80 | 0.95 | 0.95 | Pr{pathology} |
| $\text{VE}_H$ | 0.90 | 1.0 | 1.0 | Pr{severe outcomes \| pathology} |
| $\text{VE}_I$ | 0.40 | 0.80 | 0.80 | infectiousness |

**Table S5:** Aggregated VE estimates from multiple Phase III trials and other published sources (*20*). These aggregated efficacy values parameterize the generalized mRNA vaccine simulated in our ABM. Note that $\text{VE}_{SP}$ is the primary endpoint of Phase III vaccine trials.

### 3 Synthetic Population

We create a sample synthetic population of 375K people, 162K households and 34K businesses of various size using Marion County, FL, as the template for spatially explicit information, *e.g.*, geographical coordinates of locations. Marion County consists of a mixture of urban and rural areas (Fig 1b–c in Main Text) and is fairly compact, minimizing potential boundary effects.

As an overview, we first populate the space with households and the people who live in these households. We assign the individual attributes of these people according to census and survey data. Then, we determine the locations of places of interest, including workplaces, schools, long term care facilities and hospitals. Finally, we assign the daily interactions between people and the places of interests, and the interactions between households.

#### 3.1 Demographics and Households

Household placement is based on the 5-year data set (2014 to 2018) of the American Community Survey (ACS) (*21*). The US Census Bureau uses this survey to estimate the number of households in each census block group (CBG). We obtained these aggregated numbers via a shapefile provided by the Florida Geographic Data Library (*22*). Separately, the Integrated Public Use Microdata Series (IPUMS) provides fine-grained details for 5% of US households and their members (*23*) based on the ACS. Thus, for CBG $i$, we randomly sample $n_i$ households from the IPUMS dataset with replacement, where the estimate of $n_i$ is from the ACS dataset. We then randomly place the sampled households using population density maps provided by Gridded Population of the World by Socioeconomic Data and Applications Center (*24*). Specifically, we place each of the sampled households onto one of the 1 km$^2$ grids within the CBG, with probability weight proportional to the reported population density of the grids. Since all people in the IPUMS data are associated with a household, the placement of households also places people into this synthetic population. We carry over the individual attributes from the IPUMS data, including sex, age, employment and school status. As a result, household membership and age structure, as well as regular daytime activities within the synthetic population, are representative of the entire Florida population (see Figs S2 and S3).

Using data from the CDC's 2016 Behavioral Risk Factor Surveillance System (BRFSS) survey for the state of Florida (*25*), we used Bayesian mixed logistic regression to estimate the probability of a Florida resident having *at least* one COVID-19-related comorbidity given sex and age group (18 to 29, 30 to 39, 40 to 49, 50 to 59, 60 to 69, 70 to 79 and $> 79$). The comorbidities are diabetes, obesity, chronic obstructive pulmonary disease, and coronary artery disease (which includes heart attacks, coronary disease, and strokes). For every person in the synthetic population, we randomly assigned a binary comorbidity state based on the conditional probabilities we estimated.

The BRFSS survey does not cover individuals younger than 18 years old. In order to address this age-group, we assumed that obesity is the prevailing comorbidity, based on a recent study finding obesity to be a leading risk factor for COVID-19 hospitalizations among children (*26*). Though fine-grained, age-stratified data for young people in Florida are not available, nationwide data (*27*) indicate 13.9% obesity for 0 to 5 years, 18.4% for 6 to 11 and 20.6% for 12 to 17 years. These values are in concordance with less fine-grained data on Florida from the

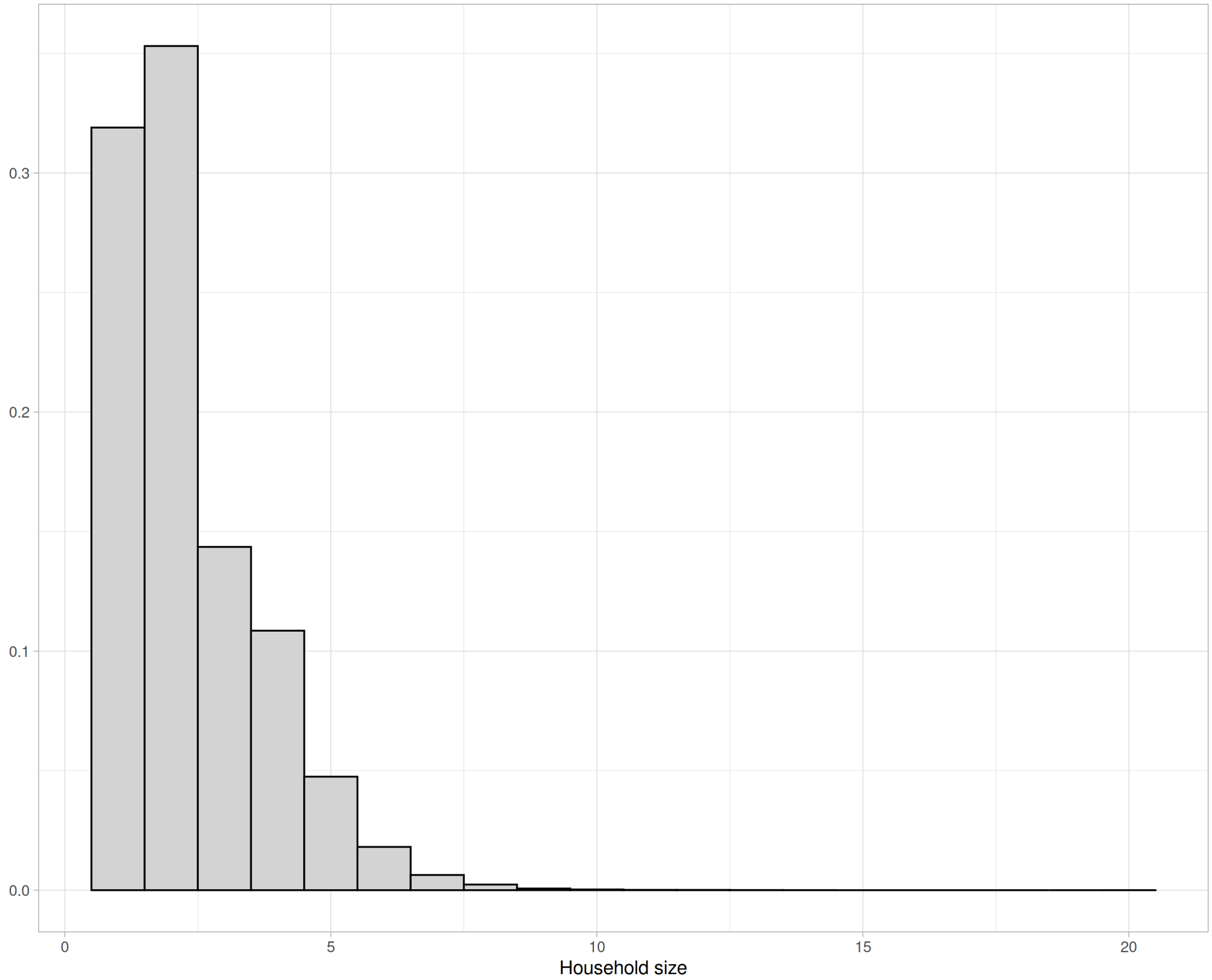


**Figure S2: Distribution of household sizes.** Households are sampled from IPUMS microcensus data for Florida. This is the resulting household size distribution for the 375K synthetic population sample.

National Survey of Children's Health (*28, 29*), which indicate 17.8% for children 10 to 17 years old and 12.7% for children ages 2 to 4 years old (enrolled in Women Infants Children Program). Therefore, we used nation-wide obesity data to represent comorbidities for children and youths.

#### 3.2 Long-term Care Facilities

Long-term care facilities (LTCFs), including nursing homes and assisted living facilities, are treated as a type of residence inhabited by multiple elderly individuals. We create these locations in the models by synthesizing two datasets. The LTCF locations and resident population totals are derived from an Agency for Health Care Administration (AHCA) dataset (*30*). The

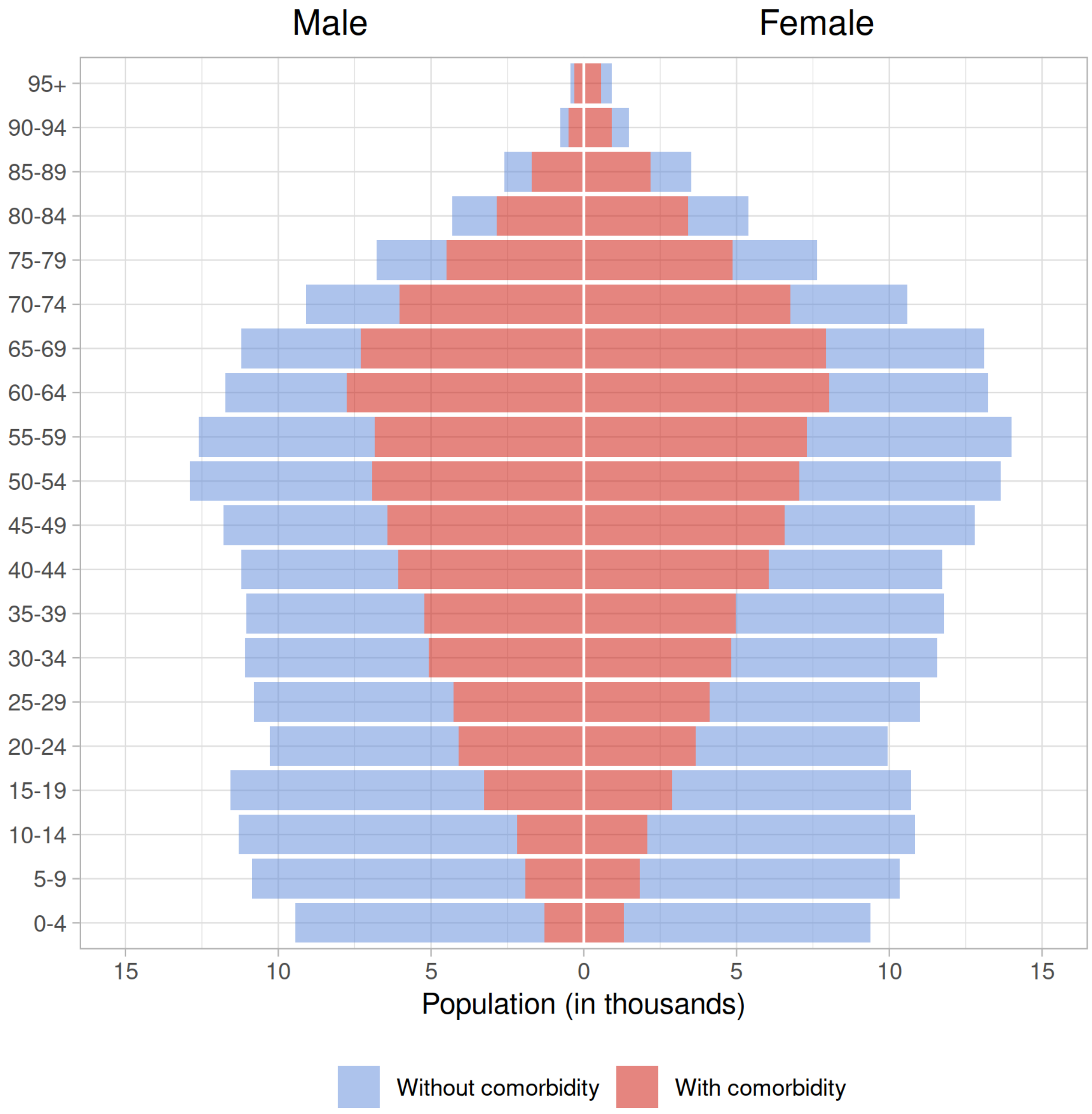


**Figure S3: Synthetic population age pyramid.** Data from the 375K synthetic population sample.

age distribution within those populations comes from the IPUMS dataset for individuals living in group quarters (*23*). However, because IPUMS "group quarters" does not distinguish by type (*e.g.*, LTCF versus correctional facility), we truncate that distribution to over 55 years of age before sampling. We do not otherwise use the 54 years of age and younger portion of the distribution.

#### 3.3 Schools

We use exact locations for schools in the synthetic population. We use a shape file created by the University of Florida GeoPlan Center with data from the National Center for Education Statistics (NCES) to locate the schools in the synthetic population. The shape file was downloaded through the Florida Geographic Data Library (*22*). Schools that are represented in this shape file include pre-kindergarten, kindergarten, elementary, secondary and high schools, as well as post-secondary institutions, *e.g.*, colleges and universities. We estimate the size of the 601 colleges and universities in Florida using *ad hoc* web searches. Our 375K synthetic population representing the spatial structure of Marion County, FL, includes 8 higher education institutions. For grade schools, we lack systematic enrollment information, and so all grade schools are treated as having the same weight for allocation purposes.

Model schools are closed during weekends and during summer breaks, based on the public school calendar for Florida schools. Transmission in schools is at times partially reduced in the model, to represent schools being open but with interventions in place, such as required masking, cohorting of students, or reducing classroom capacity.

#### 3.4 Workplaces

Workplaces in the dataset are based on the National Corporation Directory (NCD) dataset of all businesses in Marion county. We have established a data use agreement with NCD, which provides the address and the business type of all businesses in Florida. Addresses were geocoded using ArcGIS. The business type is coded using the North American Industry Classification System (NAICS), allowing us to identify essential vs non-essential businesses for purposes of modeling lockdowns. We removed schools and LTCFs from the dataset as their locations are determined using other sources. Since the sizes (number of employees) of these workplaces are not available, we developed a sampling procedure that combines (1) local data on the number of jobs from the Longitudinal Employer-Household Dynamic (LEHD) Origin-Destination Employment Statistics (LODES) Workplace Area Characteristics (WAC) dataset (*31*), and (2) nation-wide information on sizes of different types of businesses provided by the Bureau of Labor Statistics (*32*). Because the latter information consists of distributions of binned values (1 to 4, 5 to 9, 10 to 19, 20 to 49, etc) for each type of business, we first replaced this with a fitted Pareto distribution. Then, for each CBG, we sampled workplace sizes (given the distribution of local types of businesses) to yield an approximately correct number of employed persons, which was then rescaled to provide a more precise fit (see Fig S4).

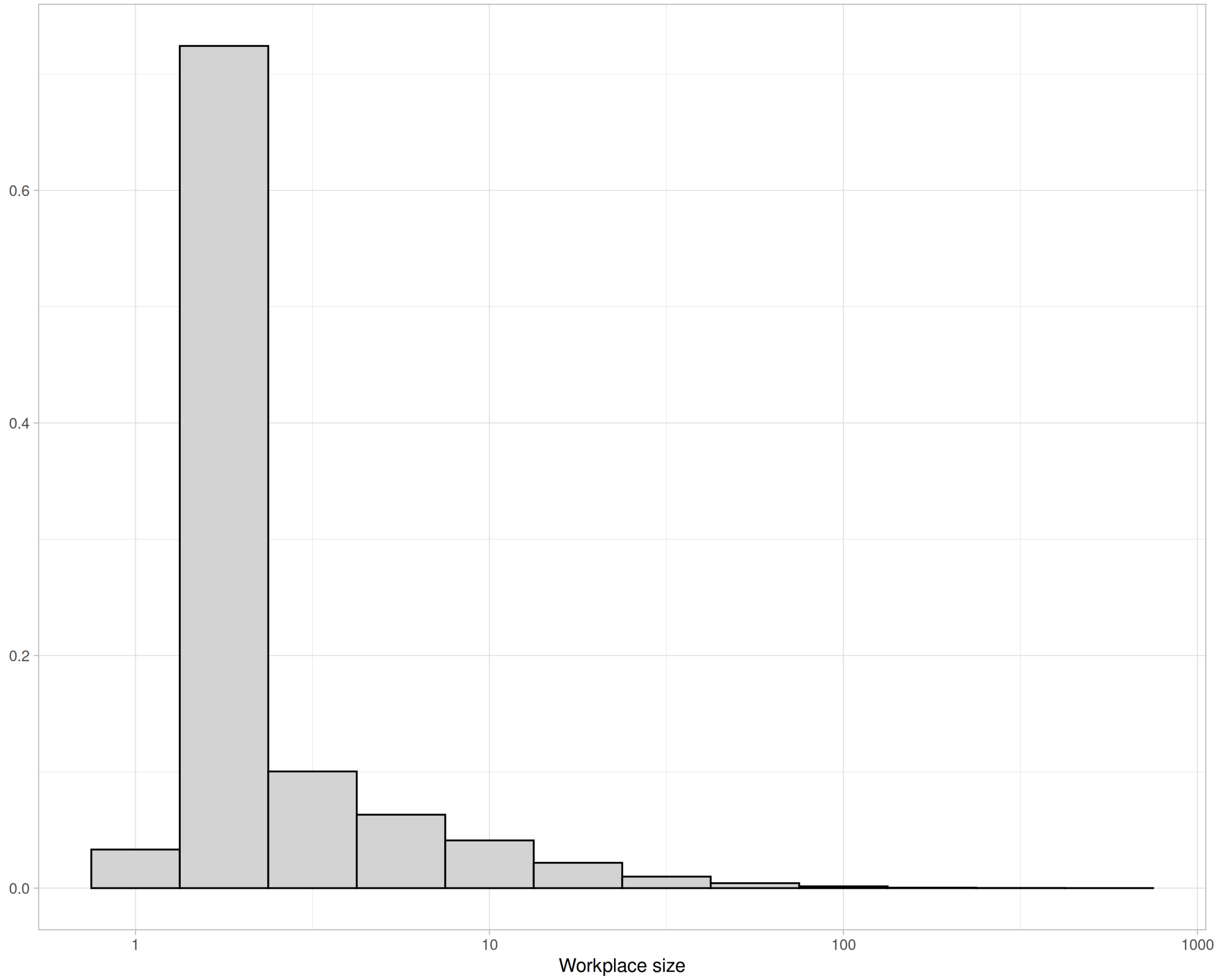


**Figure S4: Distribution of workplace sizes.** Data taken from the 375K Marion County synthetic population sample.

#### 3.5 Hospitals

Hospital locations are explicitly represented in the synthetic population based on a dataset from AHCA (*30*). Additionally, the sizes (*i.e.*, the number of beds) of these hospitals are also obtained from the dataset. We do not model hospital capacity or resource usage in this work. Hospitalized patients are removed from the transmission processes in their households, inter-household network, workplace or school, and public activity locations. If an individual's infection becomes severe, the person will probabilisitically be hospitalized; if they become critically ill, they may be admitted to an ICU. Individuals that are already hospitalized when they become critical are more likely to receive ICU care than are unhospitalized individuals who become critical. ICU care is modeled explicitly, as it reduces the mortality risk once at the critical stage (see Table S2). Additionally, the use of pharmaceutical interventions (*e.g.*, dexamethasone) further reduces the chance that death occurs while receiving ICU care (see Section 5.3).

#### 3.6 Daily Interactions Between People and Places of Interest

The synthetic population does not merely specify the locations of persons, schools, workplaces, etc, but the links between them that represent routes for movement of persons. These links are assigned using modified gravity models. For a source location $s$, the gravity model specifies a set of probabilities for choosing among a set of candidate destinations. The weight $w_i$ assigned to candidate $i$ is:

$$w_i = \frac{m_s m_i}{d_{s,i}^2},$$

where $m_s$ and $m_i$ represent the analog of mass—here the size, or capacity—that characterizes the attractiveness of $s$ and $i$, and $d_{s,i}$ is the Euclidean distance between $s$ and $i$. The model then randomly samples one entity among the candidates, with probability proportional to the weight.

Persons designated to attend school are assigned to one of the 5 geographically closest schools appropriate for the age of the person. Students 19 and older are assigned to one of the 5 nearest colleges or universities using a gravity model that incorporates enrollment size, whereas younger students are assigned randomly to one of the 5 closest age-appropriate schools (given the absence of enrollment data).

Each household of the synthetic population is randomly assigned to one of the nearest three hospitals using the gravity method, with mass equal to the number of beds in each hospital.

Individuals may be admitted to their assigned hospital if they become severely or critcally ill.

For workplaces, we first determine the workforce for schools, LTCFs, and hospitals, since they also function as workplaces. Each school is assigned 1 employee for each 7 students previously assigned. This ratio is based on assuming a teacher:student ratio of 1:14 (as per the GeoPlan school facilities dataset) and then doubling this to allow a 1:1 ratio of teachers to other employees. We assume 1 employee to 6 LTCF residents, following the Florida state requirement of minimum staffing for assisted living facilities (*33*). The number of employees in a hospital is equal to 8 times the number of occupied beds, assuming 80% occupancy, following (*34*).

Then, for a person who claimed to be employed in the ACS, we first identify at least 1000 workplaces with unfilled jobs that have the shortest straight-line distances to the person's residence. We then randomly assign the person to one of the selected workplaces based on the gravity model, with the mass equal to the number of unfilled jobs available. By starting with a large number of candidate workplaces, this method allows, *e.g.*, some people to travel long distances to larger workplaces, while generally favoring short commutes.

#### 3.7 Inter-household Network

An undirected inter-household network is also generated in order to represent possible routes of transmission via social interactions between friends, neighbors, and relatives. We first determined the number of edges for each household, which is sampled from a Poisson distribution with $\lambda$ equal to the household size. We then link households using a gravity model with an additional associativity term for households with similar levels of COVID-19 risk tolerance (see Section 3.1). Specifically, this term is $1 - |c_i - c_j|$, where $c_i$ and $c_j$ are risk tolerance thresholds assigned to households $i$ and $j$ from a uniform distribution (see Fig S5).

#### 3.8 Businesses Patronization

Each individual is associated with five businesses that are relevant to SARS-CoV-2 transmissions. This number was chosen as large enough to generate heterogeneous daily activities both between individuals and among any individual's overall time in the model, while being small

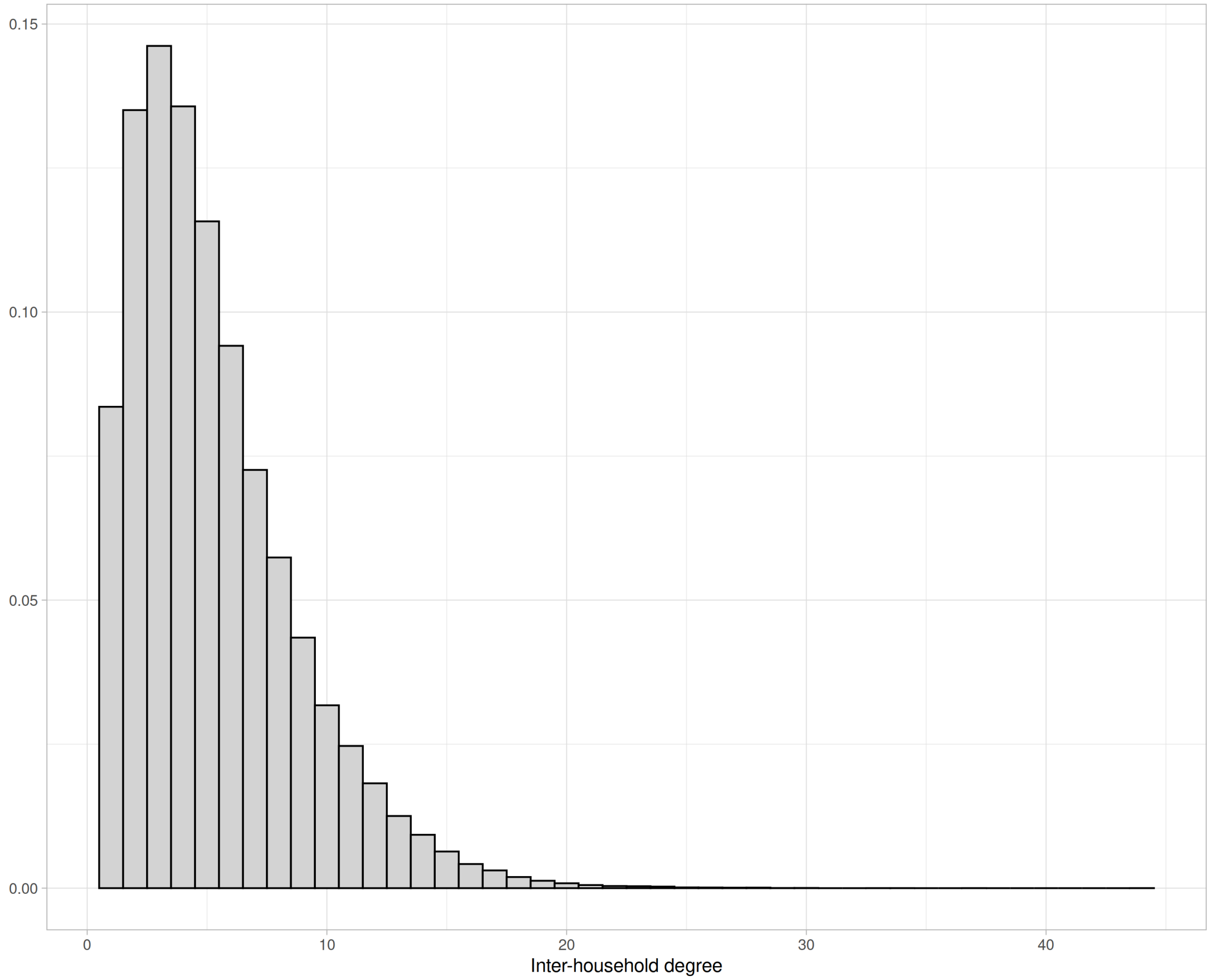


**Figure S5: Inter-household network degree distribution for the 375K synthetic population sample.**

enough to be computationally tractable for the entire population. They are allocated using a gravity model, with the "mass" equal to an assigned visitor count to these businesses. On any given day, individuals may patronize between 0 and 5 businesses in the model. The number of total hours spent patronizing businesses on a given day is Poisson(1) distributed. Each hour is equally likely to be spent at any of the 5 eligible business for that individual. The relative infectiousness of a patron compared to an employee is the number of hours spent at a business, divided by 8 (to represent an employee's typical 8 hour shift).

There are 19 types of businesses (categorized by NAICS code) that can be patronized. These NAICS business types are further categorized into two groups: high and low inherent transmission risk (see list below). Risk categories are defined using empirical data on variable infection risk by location type (*35*). All other types of businesses are assumed to have minimal in-person patronage; for these, we model only interactions between employees (see Fig S6).

High risk business types

- Full service restaurants
- Limited service restaurants
- Snack and nonalcoholic beverage bars
- Cafeterias, grill buffets, and buffets
- Drinking places (alcoholic beverages)
- Fitness and recreational sports centers
- Religious organizations

Low risk business types

- Supermarkets and other grocery (except convenience) stores
- Convenience stores
- Department stores
- Gasoline stations with convenience stores
- General merchandise stores such as hardware, sporting goods, automotive parts, pet, pharmacies, hobby and others

To determine the "mass" of each patronizable business to use in the gravity model, we use pre-pandemic visitation data. We first obtain January and February 2020 raw visitor counts of all businesses in Florida from the SafeGraph place of interest dataset (*36*). Then, for a "high risk" business in the synthetic population, we randomly select a "high risk" business in the SafeGraph dataset, and assign the latter's pre-pandemic raw visitor count to the former. We assign the visitor count for "low risk" businesses similarly.

### 4 External Introductions

Introductions happen in two phases. The population is initialized at time 0 with an exposure probability of $10^{-4}$ per person. Subsequently, individuals face an external exposure with a probability of $10^{-4}$ per day. VOCs are introduced using this mechanism, as described in Section 2.1.

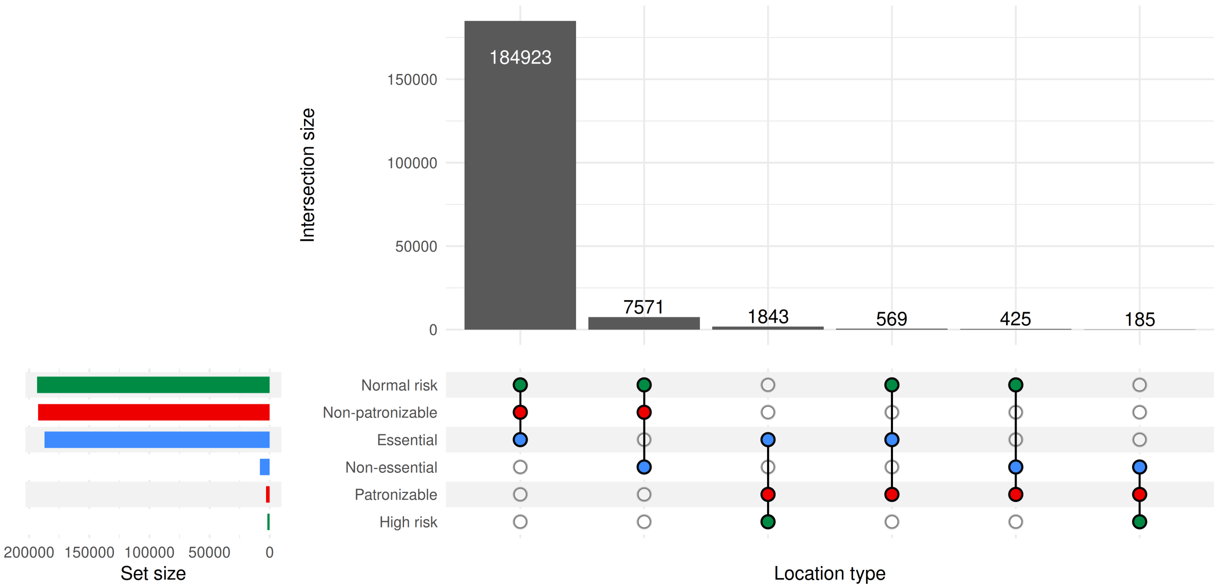


**Figure S6: Workplaces by type.** Workplaces are sorted into 3 pairs of mutually-exclusive sets: Essential/Non-essential (blue), Patronizable/Non-patronizable (red), and Normal risk/High risk (green). The matrix shows intersections of these six sets, and the bar chart shows the frequency of businesses in the intersection. Data taken from the 375K Marion County synthetic population sample.

### 5 Transmission

An individual's probability of becoming infected in a given place depends on several transmission factors (Equation S3), including the type of location ($\tau_\lambda$, see Table S6), number and infectiousness of other people at that location, and seasonality ($\psi_d$). Transmission in workplaces and schools may be partially or completely reduced via time-varying interventions ($a_d$), and workplaces may be either high or low risk ($\rho$), depending on business type. The infectiousness hazard multipliers $\omega$ of each person $i$ present at the locale on day $d$ is summed, and for some locale types is normalized by the number of other people present ($D$). These factors are multiplied to determine the transmission hazard for a susceptible person at that location on a given day, which is used to calculate the probability of exposure $T_{\lambda,d}$ for a given locale $\lambda$ on day $d$:

$$T_{\lambda,d} = 1 - e^{-a_d \psi_d \rho \tau_\lambda \sum_i \omega_{i,d}/D} \tag{S3}$$

| Location type | Value | Details |
|---|---|---|
| Household, $\tau_H$ | 0.07 | Density dependent |
| Workplace, $\tau_W$ | $\tau_H/2$ | Frequency dependent |
| Inter-household, $\tau_{IH}$ | $\tau_H/2$ | Frequency dependent |
| Health care, $\tau_{HC}$ | $\tau_H/10$ | Frequency dependent |
| Long-term care, $\tau_{LTC}$ | $\tau_H * 3/2$ | Frequency dependent |
| School, $\tau_S$ | $\tau_H/2$ | Frequency dependent |

**Table S6: Transmission hazards by location type.**

An individual's infectiousness hazard multipliers are specific to each infection they experience:

$$\omega_{i,d} = \theta_s(1 - \text{VE}_I)\phi \qquad \text{(S4)}$$

where $\theta_s$ is the infectiousness of the viral strain, relative to wildtype; $\text{VE}_I$ is vaccine efficacy against infectiousness, in the event that the individual is vaccinated; and $\phi$ is either 4 or 1/4, depending on whether the individual is high- or low-shedding. $\phi$ is determined by comparing a sampled viral load to a threshold value. For asymptomatic infections, viral load is sampled from a Weibull(6.72, 3.33) distribution, and for symptomatic infections from Weibull(7.40, 3.81) (*37*), and the deviate is compared to a threshold of 8.04. Infections above this threshold are high-shedding, and below are low-shedding. This results in approximately an 80/20 Pareto principle, where 20% of infections produce 80% of the total infectiousness.

An exposure may or may not result in infection. Susceptibility may be modulated by a range of factors such as age or immunity from vaccines and previous infections; see Section 2.2 for details.

Events in the model occur as a daily cycle of non-overlapping periods for (1) vaccinations, (2) community transmission (intra-household, work, school, inter-household), and (3) introductions. The rules for exposure differ by the locale of transmission.

**Household** A household is treated as a complete graph, such that all members are connected. The chance of exposure is $1 - (1 - \tau_H)^c$ where $\tau_H$ is a calibrated parameter representing household transmission hazard, and $c$ is the number of infected members.

**Workplace** A workplace may be closed or inactive due to interventions (see Section 5.3). The exposure hazard in a workplace is $a_d \tau_W f$ where $a_d$ is 0 if a business is closed and otherwise 1; $\tau_W$ is a calibrated parameter representing workplace transmission hazard; and $f$ is the frequency of infected individuals. Infected individuals in this setting include coworkers who are asymptomatic or mildly symptomatic and infected, non-hospitalized patrons.

**School** A school is active to some degree $a_d$ between 0 (completely closed) and 1 (fully open at pre-pandemic levels) that may differ each day due to interventions (see Section 5.3). The chance of exposure is $a_d \tau_S f$ where $\tau_S$ is a calibrated parameter representing the school transmission hazard, and $f$ is the frequency of infected individuals. Infected individuals in this setting include infected students and employees who are asymptomatic or mildly symptomatic.

**Inter-household** People may interact with others in their inter-household network (see Section 3.7). If households interact on a given day, the exposure hazard is $\tau_{IH} f$ where $\tau_{IH}$ is a fitted parameter representing the inter-household transmission hazard, and $f$ is the frequency of infected individuals. Infected individuals in this setting include infected, non-hospitalized persons in the other household.

**Healthcare Facilities** The chance of exposure is $\tau_{HC} f$ where $\tau_{HC}$ is a calibrated parameter representing transmissibility within healthcare facilities, and $f$ is the frequency of infected individuals. Infected individuals in this setting include patients and infected employees who are asymptomatic or mildly symptomatic.

**Long-term Care Facilities (LTCFs)** The chance of exposure is $\tau_{LTC} f$ where $\tau_{LTC}$ is a calibrated parameter representing transmissibility within LTCFs, and $f$ is the frequency of infected individuals. Infected individuals in this setting include all infected residents, and infected employees who are asymptomatic or mildly symptomatic.

**Public** Public transmission is captured within workplace transmission (described above) via customer patronage, which varies by day. For details on how people select where to spend time outside of their home or workplace, see Section 3.8.

#### 5.1 Detection and Reporting

Transmission dynamics in the model are driven principally by infection dynamics, but it is not possible to directly compare infections in the model with empirical data. Particularly for asymptomatic/mild infections and infections early in the pandemic, the ascertainment probability is likely low. In addition, empirical data typically associates a case with a report date, and not, *e.g.*, when that infection began, or when the sample was collected. The time lags from detection to report have varied during the course of the pandemic, and differ between deaths and non-fatal cases.

For purposes of making comparisons between the model and empirical data, we superimpose a detection and reporting model on the biological model of transmission. This is intended to reflect aspects of real-world testing, diagnosis and reporting lags. Whether an infection is reported depends on symptom severity as well as when it occurs (*e.g.*, early or late in the modeled time period). Parameters related to the observation process are based on line-list and all-cause mortality data and include: (1) COVID-19 death ascertainment, (2) case fatality ratios, and (3) reporting lags.

We assume that the probability of detection increases monotonically with outcome severity. A given infection may be first detected at any stage, and once it is detected, it remains detected, *e.g.*, if an infection is detected while asymptomatic, and then the individual dies, this is counted as a known death and the death is reported with some lag (see Section 5.1.1). In the model, the chances of detection are treated as time-dependent (Fig 3 in Main Text). Note that an overall chance of detection cannot be determined from these parameters alone as it depends on the distribution of infection outcomes, which changes throughout a simulation.

##### 5.1.1 Reporting Lags

The observation model includes a time-dependent delay in reporting based on observed changes in reporting. For the state of Florida, the reporting delay decreased from a median of over 12 days early in the epidemic to a median of 1.5 days by August 2020. In the model, the reporting delay for a case is sampled from a gamma distribution with a shape that reflects the actual reporting delays for Florida.

#### 5.2 Contact Tracing

Some vaccination strategies and NPIs (*i.e.*, ring vaccination and quarantining, respectively) depend on daily contact tracing to identify targets of these interventions. Contact tracing is performed each simulated day after all transmission events are modeled. Index cases are identified from all newly detected infections with a probability of 70% (assumed value). For each index case, a location-specific number contacts are calculated following the rules described in Table S7. Then, the calculated number of contacts is sampled from the corresponding location (*e.g.*, the index case's home, neighbors, workplace/school). Interactions from public activities are not considered as contact-generating. This contact-detection process is repeated to the desired depth; for this work, contact tracing was performed to a depth of two (*i.e.*, identifying contacts and contacts-of-contacts of index cases).

| Contact type | Number of contacts traced |
|---|---|
| Household member | All members |
| Nursing home residents | Poisson(5) |
| Inter-household contact | Poisson(5) |
| Coworker | Poisson(3) |
| Schoolmate | Poisson(3) |

**Table S7: Rules for selecting number of contacts by location type.** Expected numbers of traced contacts in each location are assumed values.

#### 5.3 Time-dependent Interventions

**School Closures** We model school closures directly with schools fully open at the start of the simulation. On 15 March 2020 schools close, stay closed until half-opening on 31 August 2020 for the first full school year of the pandemic, ending 16 June 2021. Schools then close for the summer and reopen at 80%, starting 09 August 2021. See Section 5 for details on how school activity affects transmission.

**Nonessential Business Closures** The coding of business type (see Section 3.4) allows us to identify essential vs non-essential businesses as directed by Florida Executive Order 20-91 (*38*). As a consequence of this order, non-essential businesses closed between 03 April 2020 and 04 May 2020. In the model, the effect of the order is that non-essential business are removed from the list of possible sites of transmission, thus workers at essential businesses continue to be exposed, and workers at non-essential business face no exposure from their assigned work location.

**Dexamethasone** Modeled as a 35% reduction in ICU mortality. This effect is phased in logistically, with an inflection point on 15 June 2020, and a slope of 1.

#### 5.4 Personal Protective Behaviors

Individuals during the pandemic made a wide range of behavioral changes to reduce their chance of SARS-CoV-2 infection, including masking, physical distancing, hand-washing, switching to outdoor activities, etc. Adoption of these measures was heterogeneous across populations, changed over time, and varied in effectiveness. Parameterizing a personal protective behavior (PPB) model directly from data has proven difficult, but omitting behavioral changes altogether, in our experience, has made it impossible to reproduce COVID-19 trends.

We represent the impact of all PPBs in terms of individuals dynamically reducing their risk of exposure by avoiding inter-household social interactions (see Section 3.7) and high-transmission-hazard businesses (see Section 3.8) like restaurants and gyms. These decisions are made at the household level, and in the case of inter-household interactions, either household in a dyad can break contact.

In the model, a household's decision to adopt PPBs is determined by comparing two quantities: (1) a fixed risk tolerance threshold, sampled from Uniform(0,1) during the synthetic population generation phase, and (2) a dynamic societal (population-wide) risk perception level, on [0, 1]. The latter is represented as a sequence of fitted values ("anchor points"), spaced two weeks apart for the duration of the simulation, with linearly interpolated values between the fitted values. In brief, the fitting procedure involves running the simulation in overlapping, 8-week intervals, and adjusting the most recent anchor point until the simulated number of cumulative reported cases is in agreement with empirical data; full details can be found in Section 7.

### 6 Vaccination

A description of the vaccination distribution strategies we consider can be found in the Main Text Methods, under Scenarios, with a recap in Section 6.2. The strategies essentially differ in how individuals are prioritized for vaccination. Below, we focus on the mechanics of how vaccines are administered in the model.

#### 6.1 Distribution System Structure

At the start of a simulation, a dose availability time series is read in, specifying the number of doses that can potentially be distributed on each day. Any doses that are not distributed are rolled over to the following day. Elements in this time series are decremented as doses are consumed.

A vaccine-eligible pool (mathematically, a set) is maintained for the duration of the simulation. If, on a given day, one or more doses are available according to the time series, and one or more people are in the pool, a random individual is chosen from the pool for vaccination. Sampled individuals are always removed from the pool, as they either are vaccinated (in which case a dose is consumed) or have died since being queued (no dose consumed). This process continues until either the pool is empty, or the number of doses for that day reaches zero.

When people receive a first dose, they are staged for revaccination with a second dose. On the day they become eligible for revaccination, they are reinserted into the vaccine-eligible pool: the strategies we consider here, therefore, do not attempt to explicitly prioritize based on dose ordinality. The same process repeats for subsequent doses. We observe, nonetheless, that some prioritization occurs as a result of other criteria for being inserted into the pool; see Figs S1 and S2 in S2 Additional Results.

The description above is algorithmically equivalent to how this system works; the actual implementation is somewhat more complex, as it allows for additional prioritization criteria not used here.

#### 6.2 First Dose Delivery

All campaigns use the same general blueprint for administering first doses to people, but the four campaign strategies we consider differ in how individuals are placed in the eligibility pool:

**Mass Vaccination** The standard mass vaccination campaign places all vaccine-elgible (*i.e.*, $\geq 5$ years of age) people in the pool at the start of the simulation. They remain there until doses become available.

**Ring Vaccination** On each day that the campaign is active, detected index cases, their contacts, and their contacts-of-contacts are probabilistically traced. Any unvaccinated index cases or contacts are then scheduled to be eligible for first doses on the same day they were traced.

**Risk-prioritized Vaccination** On the first day of the simulation, all age-eligible people are sorted by their risk of severe disease given infection with the wildtype strain of SARS-CoV-2 (see Table S8). After sorting, deciles are constructed. The highest-risk decile is placed in the pool for vaccine eligibility immediately. Once all members of this decile group receives first doses, the next highest decile group becomes eligible. This repeats as each group's members receive first doses.

| Age bin | Risk without comorbidity | Risk with comorbidity |
|---|---|---|
| 0–9 | 0.011 | 0.065 |
| 10–19 | 0.0048 | 0.031 |
| 20–29 | 0.0073 | 0.047 |
| 30–39 | 0.015 | 0.080 |
| 40–49 | 0.026 | 0.12 |
| 50–59 | 0.047 | 0.18 |
| 60–69 | 0.097 | 0.31 |
| 70–79 | 0.19 | 0.45 |
| 80+ | 0.21 | 0.43 |

**Table S8: Probability of severe disease given wildtype infection.**

**Age-prioritized Vaccination** This strategy is nearly identical to the risk-prioritized vaccination strategy except that people are sorted by age instead of actual risk value.

##### 6.3 Revaccinations

All distribution strategies use the same mechanisms to control revaccinations. As described above, when individuals are vaccinated, they may be staged for revaccination at some point in the future. In this work, we consider a three-dose regimen, with intervals of 28 days between the first and second doses and 240 days between the second and third doses (reflective of the median lag in second and third dose administration in Florida). Revaccinations use the same pooling and random selection process as described for first doses.

#### 7 Model Calibration

We calibrated the model through an iterative process of manually adjusting parameters that are not well established in the literature, (such as the probability that a critically ill individual will seek and receive intensive care and the probability mild cases were reported during the omicron wave in Florida) followed by algorithmically fitting the societal risk perception (SRP) time series used by the PPB model. See Section 5.4 for details on the PPB model itself.

The SRP time series was fit against cumulative reported cases. As described previously, SRP values are represented as "anchor points" spaced 2 weeks apart, with daily values interpolated linearly. Anchor points are chosen in succession from the beginning to the end of the period for which we have empirical COVID-19 case data (*i.e.*, through 27 June 2022) using a heuristic, greedy algorithm. The algorithm simulates an 8-week window that shifts 2-weeks forward each time an anchor point value is accepted. The anchor point being fitted is the one two-weeks into the window. At the start of the simulation, the first two SRP anchor points are constrained to have the same value, which is initialized at 0, although the procedure is robust to initial value assumptions. Subsequently, the first two SRP anchor points within a window will tend to differ, with linearly interpolated values in between, while the next 6 weeks (tentatively) have the same value as that of the anchor point being fitted.

Goodness of fit is evaluated as the sum of weighted differences between simulated and empirical cumulative reported cases that is then normalized by the 3-day moving average in empirical reported cases centered on the anchor point:

$$G = \frac{1}{\mu_{emp,d+13:d+15}} \sum_{i=d}^{d+56} w_i (CRC_{sim,i} - CRC_{emp,i}) \tag{S5}$$

where $\mu_{emp,d+13:d+15}$ is the 3-day mean of empirical reported cases, centered around the anchor point on day $d$; $w_i$ is the weight associated with the day that is $i$ days into the 8-week fitting window; and $CRC_{sim,i}$ and $CRC_{emp,i}$ are the cumulative reported cases as of day $i$ for the simulated and empirical data, respectively. Normalizing by the 3-day mean is needed in order to use a constant value for the acceptance threshold. We use a value of 20 for the acceptance threshold; values much greater than this resulted in fits that diverged and could not recover due to the greedy nature of the algorithm, while smaller values simply take longer to run. If the acceptance criterion was not satisfied (*i.e.*, too many or too few simulated reported cases were produced), a new anchor point value was chosen using a binary search on [0,1]. The 8-week window is then re-simulated with the new anchor value and goodness of fit is re-evaluated.

This process is repeated until the the fit score is below the acceptance threshold, or the difference between the previous and next proposed anchor value is 0.01 (so that the search is guaranteed to end). Overfitting was avoided in two ways: (1) if the sign of the error changed unexpectedly while fitting a given anchor point, *i.e.*, SRP was increased, but cases did not decrease, fitting for that point was halted, and the best value that had been seen thus far was chosen; and (2) for the results presented in this work, we used the mean SRP curve of 1000 fitted curves, each simulated using different PRNG seed and our Florida reference scenario.

Statistically, this approach is using the PPB model to account for residuals. Because it only changes two kinds of interactions (patronage of high-risk businesses, and inter-household contacts) by a limited amount, however, it can only make a reasonable model better, and not salvage a bad one. We further believe that the resulting PPB model is credible, because this procedure consistently produces a societal risk perception curve with peaks that lag behind reported cases, and with peaks that tend to sequentially decrease in height. This matches our intuition (and informal first-hand experience) that people reacted to information that was somewhat old, and that the strength of that reaction decayed due to fatigue.

We evaluated the fit of the PPB model as follows: If the fitted SRP curves had any fitted anchor point values of 0 or 1, we took this to mean that the baseline transmission hazard was too low or too high (respectively), adjusted the baseline hazard manually and repeated the fit. If the SRP fitting procedure was able to reproduce the cumulative reported case curve but seroprevalence was off, we adjusted the case reporting probabilities. If the SRP fit was successful but cumulative hospitalizations and/or deaths were off, we adjusted the probability that severe infections resulted in hospitalizations. If metrics were generally reproduced well, but failed during a particular wave (typically delta or omicron), we adjusted VOC-specific parameters (*e.g.*, pathogenicity) as long as those parameters were still within the range of published estimates.

### 8 Simulations and Analysis of Outcomes

We simulate the pandemic from 10 February 2020 to 23 April 2022. For the main text results, we recorded the weekly number of infections and deaths of each simulation for analysis. For each scenario, we calculated the median ($N = 1000$) weekly numbers of infections and deaths. When calculating relative performance between scenarios, replicates are matched by random number seed before calculating ratios or differences. Using scenarios with no quarantining and a standard mass vaccination program as control, we use the median values to calculate the cumulative effectiveness and cumulative infections and deaths averted at time $t$ using following formulas:

$$\text{Cumu. outcomes averted} = C_{\text{control},t} - C_{\text{scenario},t}$$
$$\text{Cumu. effectiveness} = 1 - \frac{C^*_{\text{scenario},t}}{C^*_{\text{control},t}}$$

where $C_{\cdot,t}$ is the cumulative number of cases or death at time $t$ and $C^*_{\cdot,t}$ is the cumulative number of cases or death since the scenario-specific intervention started. Because simulation replicates are matched by PRNG seed, outcomes averted and effectiveness are always 0 and 1, respectively, prior to the start of interventions.

### 9 Data Management, Software and Execution

Using ESRI shapefiles and CSV files as input, we use customized scripts in R with packages such as `tidyverse`, `sf`, `raster` and `brms` to generate the synthetic population (see Section 3), as well as to perform analysis and visualization of simulated data. We use R and its interface with C++ (*i.e.*, `Rcpp` package) to implement the gravity method described in Section 3.6. We store the synthetic population in a SQLite database using `RSQLite` package in R.

We compile the model using GCC (*39*), importing GSL (*40*) for pseudo-random number generation, and perform simulations on the University of Florida High Performance Computing Cluster. On an Intel Xeon E5-2683 v3 @ 2.00GHz, each simulation takes ∼2.8 minutes per simulated year, and has a memory footprint of ∼400 MB for a population of 375k. Run time and memory usage are approximately linear with population size.

The C++ simulation source code is available at `https://github.com/tjhladish/covid-abm`. R code to analyze simulation results, and generate result figures is also available in that repository. The R source code for generating the synthetic population is available at `https://github.com/kokbent/synthpop-fl`.

30. Florida Agency for Health Care, Facility/Provider Locator.

31. US Census Bureau, LEHD Origin-Destination Employment Statistics Data (2002-2019) [computer file].

32. US Bureau of Labor Statistics, QCEW Data Files.

33. Florida Department of State, 58A-5 : Assisted Living Facilities.

34. Statista, Hospital staffing ratio per bed by MHS ownership U.S. 2014.

35. S. Chang, *et al.*, *Nature* **589**, 82 (2021).

36. SafeGraph Data for Academics.

37. P. Z. Chen, *et al.*, *eLife* **10**, e65774 (2021).

38. State of Florida Office of the Governor Executive Order Numner 20-91 (Essential Services and Activities During COVID-19 Emergency) (2020).

39. GCC, the GNU Compiler Collection - GNU Project.

40. M. Galassi, ed., *GNU scientific library reference manual: for GSL version 1.12* (Network Theory, Bristol, 2009), third edn.
